# Histopathology-based Protein Multiplex Generation using Deep Learning

**DOI:** 10.1101/2024.01.26.24301803

**Authors:** Sonali Andani, Boqi Chen, Joanna Ficek-Pascual, Simon Heinke, Ruben Casanova, Bernard Hild, Bettina Sobottka, Bernd Bodenmiller, Tumor Profiler Consortium, Viktor H. Koelzer, Gunnar Rätsch

## Abstract

Multiplexed protein imaging offers valuable insights into interactions between tumors and their surrounding tumor microenvironment (TME), but its widespread use is limited by cost, time, and tissue availability. We present HistoPlexer, a deep learning framework that generates spatially resolved protein multiplexes directly from standard hematoxylin and eosin (H&E) histopathology images. HistoPlexer jointly predicts multiple tumor and immune markers using a conditional generative adversarial architecture with custom loss functions designed to ensure pixel- and embedding-level similarity while mitigating slice-to-slice variations. A comprehensive evaluation on metastatic melanoma samples demonstrates that HistoPlexer-generated protein maps closely resemble real maps, as validated by expert assessment. They preserve crucial biological relationships by capturing spatial co-localization patterns among proteins. The spatial distribution of immune infiltration from HistoPlexer-generated protein multiplex enables stratification of tumors into immune subtypes. In an independent cohort, integration of HistoPlexer-derived features into predictive models enhances performance in survival prediction and immune subtype classification compared to models using H&E features alone. To assess broader applicability, we benchmarked HistoPlexer on publicly available pixel-aligned datasets from different cancer types. In all settings, HistoPlexer consistently outperformed baseline methods, demonstrating robustness across diverse tissue types and imaging conditions. By enabling whole-slide protein multiplex generation from routine H&E images, HistoPlexer offers a cost- and time-efficient approach to tumor microenvironment characterization with strong potential to advance precision oncology.

## 1 Introduction

Tumors are complex systems that acquire hallmark traits by establishing a supportive tumor microenvironment (TME) which facilitates tumorigenesis and metastasis [1, 2]. Understanding cancer cell interactions with surrounding tissue provides critical insights into disease progression and therapeutic response [3–5]. Multiplexed immunohistochemistry and immunofluorescence (mIHC/IF) technologies, such as Imaging Mass Cytometry (IMC), enable spatially-resolved quantification of up to 40 protein markers, offering comprehensive visualization of tumor-TME interactions [4, 6, 7]. These technologies facilitate analysis of spatial cell distribution, phenotype co-localization, and cellular community interactions—factors recognized as valuable for clinical decision-making [4, 5, 8, 9]. However, IMC is limited by low throughput, high cost, and coverage restricted to small Region-of-Interests (RoIs), hindering its broader clinical adoption.

In contrast, Hematoxylin & Eosin (H&E) staining remains the gold standard for cancer diagnosis in clinical practice due to its low cost, high throughput, and coverage of entire tissue sections. H&E images reveal crucial morphological features of tissue organization that aid in cancer grading, proliferation assessment, and staging [10]. Recent advances in Deep Learning (DL) have shown that these features can inform the prediction of protein markers. Several studies have successfully predicted single markers such as pan-cytokeratin for pancreatic cancer [11], HER2 for breast cancer [12], and Ki-67 for neuroendocrine and breast cancers [13] directly from H&E images. One study has predicted two markers, specifically one tumor and one immune marker [14]. Only a few studies have attempted to predict a diverse set of protein markers, with a focus, however, solely on either tumor [15, 16] or immune markers [17], limiting their utility for investigation of tumor-TME interactions. Additionally, these studies either employ separate models for each marker [15, 17] or lack quantitative validation on the advantages of multiplexed prediction with a single model [14, 16, 17].

To address these limitations, we introduce HistoPlexer, a DL model that generates comprehensive protein multiplexes from H&E images. HistoPlexer simultaneously predicts 11 markers, consisting of both tumor and immune markers, enabling an integrative visualization of tumor-host interactions. We train HistoPlexer on metastatic samples from the Tumor Profiler Study (TuPro) [18] using paired H&E and IMC images from serial sections. Through rigorous quantitative evaluation, we demonstrate the importance of simultaneous marker prediction through improved model performance and enhanced spatial co-localization of markers. We validate the biological relevance of generated IMC images through cell-typing and immune phenotyping analysis, particularly in characterizing immune-hot (inflamed) and immune-cold (excluded/desert) tumors based on CD8+ T-cell distributions. We also demonstrate the generalizability of the trained HistoPlexer model on an independent patient cohort from the human skin cutaneous melanoma (SKCM) study of The Cancer Genome Atlas (TCGA) project [19]. Additionally, we showcase the generalizability of our approach through experiments conducted on two publicly available multiplexed datasets [14, 20], further validating its adaptability across different cancer types.

Our results demonstrate that HistoPlexer generates high-quality IMC images that closely align with real data distributions. The generated multiplexes enable precise immune phenotyping through spatial analysis of tumor-immune cell interactions, particularly in distinguishing immune-hot and cold subtypes. We show that simultaneously predicting multiple protein markers preserves biologically meaningful relationships among them. Furthermore, by augmenting H&E Whole-Slide Images (WSIs) with generated IMC multiplex, HistoPlexer improves both survival and immune subtype prediction on the TCGA-SKCM dataset, indicating its potential for enhancing clinical decision-making in precision oncology.

## 2 Results

### 2.1 HistoPlexer: a toolkit for histopathology-based protein multiplex generation

HistoPlexer is a generative model based on conditional GAN (cGAN) that predicts spatially-resolved profiles of multiple proteins simultaneously from a single input H&E image. The model is trained on paired H&E and multiplexed IMC images (Fig. 1a) extracted from aligned H&E and IMC RoIs. During training, the H&E images are input to the *translator* G, which learns to generate protein multiplexes (*i.e*., IMC images) based on the tissue morphology from high-resolution H&E images. The *discriminator* D receives the generated IMC and input H&E images and scores their similarity to ground-truth IMC images (Fig. 1b(i)). The translator and discriminator are trained adversarially using a least squares Generative Adversarial Network (GAN) loss, such that the generated IMC images can deceive the discriminator into classifying them as real. In addition to the GAN loss, we incorporate two additional losses to ensure both pixel-level and patch-level consistency between the generated and ground truth (GT) IMC images. The pixel-level consistency loss calculates the *L*_1_ distance between the generated and GT IMC images. However, since the H&E and GT IMC images are obtained from serial sections of the tissue block, a degree of spatial displacement in tissue organization exists between consecutive slices (termed slice-to-slice variation). Although aligned at the structural level via template matching, consecutive slides from real-world diagnostic material are not pixel-level aligned. To account for these differences, we adopt the Gaussian Pyramid loss [12], which relaxes the alignment constraint by evaluating the similarity between the generated and GT IMC images at multiple scales (Fig. 1b(ii)). For patch-level consistency, we utilize a patch-wise contrastive loss to ensure that corresponding patches in the generated and GT IMC images are closer in the embedding space than distant ones (Fig. 1b(iii)). We also incorporate adaptive weights for different patches based on their proximity to GT, as described in [21].

**Fig. 1:**
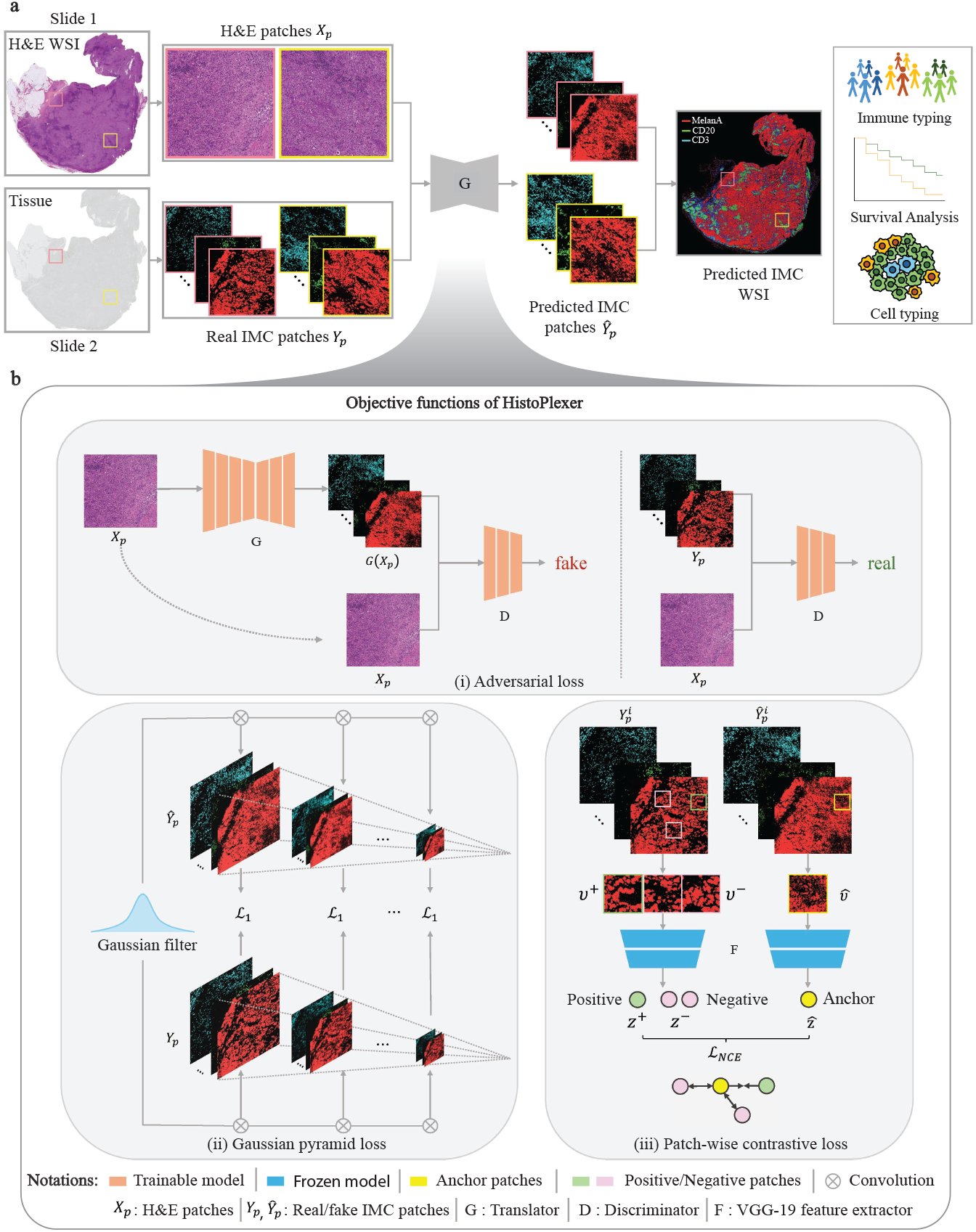
Overview of HistoPlexer architecture. **a** The HistoPlexer consists of a translator G that takes H&E and IMC images as input and predicts protein multiplexes from morphology information encoded in the H&E images, ultimately generating protein multiplex on the WSI level from H&E input. **b** The objective functions of HistoPlexer contain the GAN adversarial loss, gaussian pyramid loss with average L1 score across scales and patch-wise contrastive loss with anchor from generated IMC and positive and negative from GT IMC.

We build HistoPlexer framework using a multimodal metastatic melanoma dataset generated by the Tumor Profiler Study [18]. Each patient was characterized by multiple modalities, including H&E and IMC images. RoIs of 1 mm^2^ were selected on each H&E WSI based on visual inspection by a pathology expert and IMC data was generated for those RoIs on a consecutive section of the same tumor block. Using template matching [22], we created a paired dataset of 336 H&E and IMC RoIs from 78 patients. We focus on predicting 11 protein markers that are essential for characterizing the tumor and its surrounding TME. These include tumor markers (MelanA, S100, gp100, SOX10), immune markers (CD3, CD8a, CD20, CD16, CD31), and antigen-presentation markers (HLA-ABC, HLA-DR).

### 2.2 HistoPlexer generates accurate and realistic protein multiplexes

We benchmark HistoPlexer against Pix2pix [23] and PyramidP2P [12], evaluating each method in two settings: multiplex (MP) and singleplex (SP). In the MP setting, a single model is trained to predict all markers simultaneously, whereas in the SP setting, separate models are trained to predict each marker individually, and the predictions are subsequently stacked to form a (pseudo-)multiplexed output. All models are trained on 231 and tested on 105 RoIs.

We evaluate the quality of generated IMC images using Multiscale Structural Similarity Index (MS-SSIM) [24] to measure perceptual similarity across multiple scales, Root Mean Squared Error using Sliding Window (RMSE-SW) to quantify pixel-level differences [25] and Peak Signal-to-Noise Ratio (PSNR) [26] to assess overall signal fidelity by measuring pixel-level distortions. Our results show that the HistoPlexer model trained in the MP setting achieves the highest MS-SSIM and PSNR values and lowest RMSE-SW (Fig. 2a), suggesting higher similarity to GT IMC images generated from consecutive tissue sections. Additionally, models in the MP setting consistently outperform those in the SP setting across all methods, indicating that simultaneous prediction of all markers improves performance by capturing inter-marker correlations. The performance of individual markers for the HistoPlexer-MP model is detailed in Extended Data Table 4. For completeness, we have also included a comparison with CycleGAN [27] in Extended Data Table 3. The results on pixel-aligned DeepLIIF datasets [14, 20] have been included in Section S1.1.2.

**Fig. 2:**
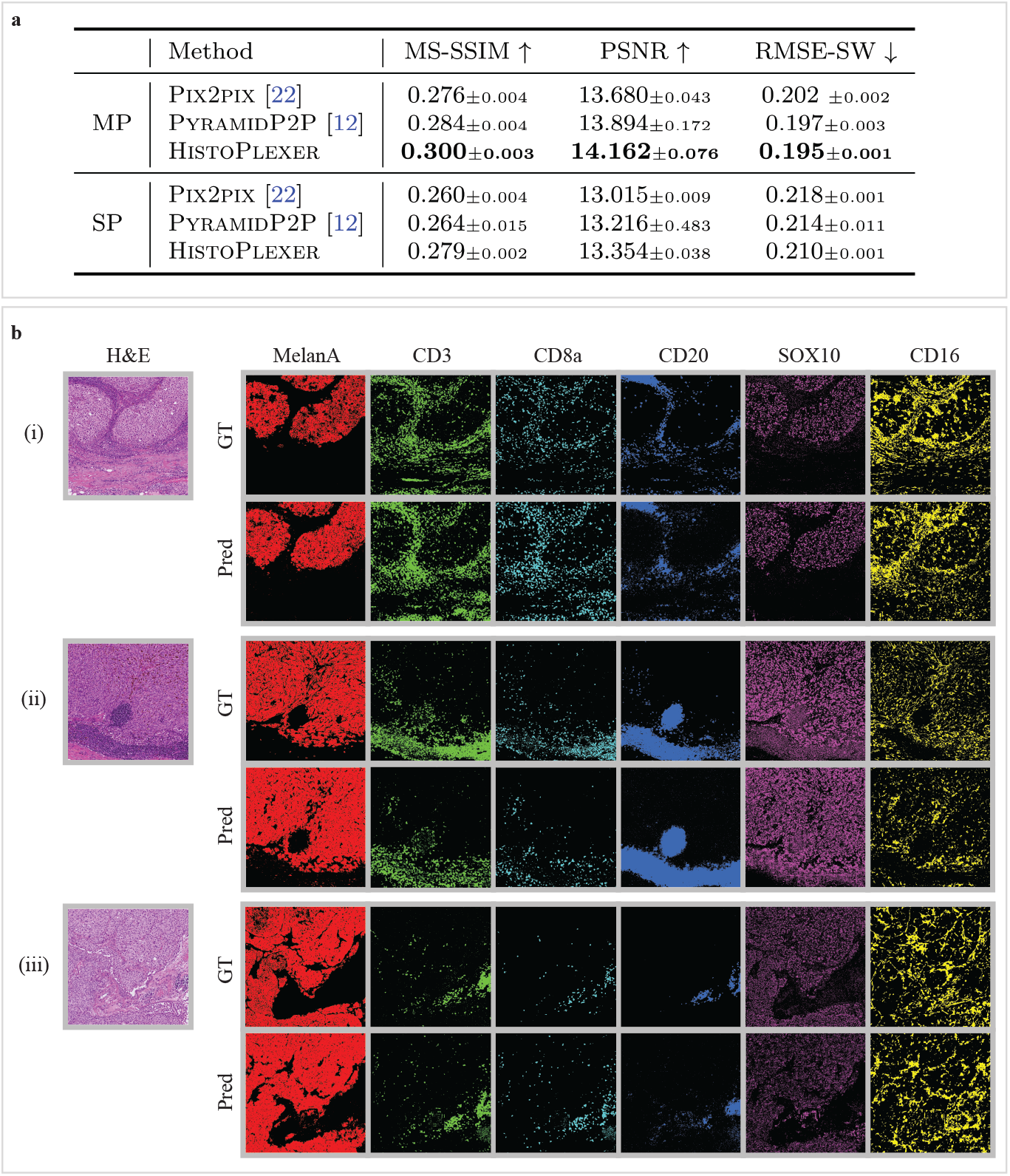
RoI-level assessment of HistoPlexer. **a** Quantitative assessment and comparison against benchmarks using MS-SSIM, PSNR and RMSE-SW for multiplex (MP) and singleplex (SP) settings. ↑ arrow indicates higher values are better. ↓ arrow indicates higher values are better. Best results are highlighted in **bold. b** Qualitative assessment of HistoPlexer for three RoIs. H&E as input to HistoPlexer (first column) and expression profiles of individual markers: MelanA, CD3, CD8a, CD20, SOX10 and CD16 (from second to last column). Top row: ground-truth (GT) expression profiles from IMC modality; bottom row: predicted (Pred) expression profiles from Histo-Plexer.

We qualitatively evaluate the generated IMC images by comparing them with the GT (Fig. 2b and Extended Data Fig. 1) and observe strong alignment in global patterns. However, pixel-level correspondence is not expected due to the inherent slice-to-slice variations. In some instances, we observe confusion between CD20 and the CD3/CD8a markers. For instance, in the bottom-right region of Fig. 2b (ii), CD20 is overexpressed while CD3 and CD8a are underexpressed. This may result from the visually similar morphology of B- and T-cells in H&E images, leading to confusion between their markers (CD20 for B-cells and CD3/CD8a for T-cells) [28].

To evaluate the perceived realism of generated IMC images, we use the Human Eye Perceptual Evaluation (HYPE) framework [29], in which experts assess real versus generated IMC images for specific markers shown alongside corresponding H&E images. Since H&E staining reveals critical morphological patterns for identifying tumor regions and lymphocytes [28], we constructed two evaluation sets: tumor-associated markers (MelanA, S100, gp100, SOX10) and lymphocyte markers (CD20, CD3, CD8a). For each set, two experts independently reviewed 250 image pairs, evenly split between real and generated, sampled from test RoIs and augmented through small translations and rotations. The evaluation yields mean HYPE scores of 41.8%(*±*0.3%) for lymphocyte markers and 42.8%(*±*0.6%) for tumor markers. The generated images achieved HYPE scores of 61.6% (*±*1.3%) and 72.8% (*±*1.1%), indicating that the majority (*>*50%) were perceived as real by domain experts, thereby demonstrating high visual realism.

Next, we go beyond pixel-level evaluation by identifying relevant cell types. We use GT cell-type annotations from the GT IMC training set, following [8], and train a Random Forest classifier [30] based on average marker expression per cell to classify them into five classes: tumor cells, B-cells, CD8+ T-cells, CD4+ T-cells, and others. This classifier is then applied to both GT and generated IMC images in the test set to obtain cell-type maps (Fig. 3a). We visualize RoIs from the tumor center and the tumor front at the tumor–TME interface and examine spatial patterns based on immune subtype labels. We observe that immune “hot” tumors, characterized by high immune cell infiltration, show strong interactions between tumor and CD8+ T-cells (Fig.3a(i)), whereas immune “cold” tumors, with low immune presence, display minimal immune cell interaction, especially in the tumor center (Fig.3a(ii)). Immune “cold” RoIs at the tumor front likewise exhibit sparse or clustered immune cells with minimal interaction with tumor cells (Fig.3a(iii), (iv), (v)). The strong alignment between predicted and GT cell-type maps, as well as their spatial organization, suggests that HistoPlexer effectively captures morphological features in H&E images that are relevant for predicting cell types.

**Fig. 3:**
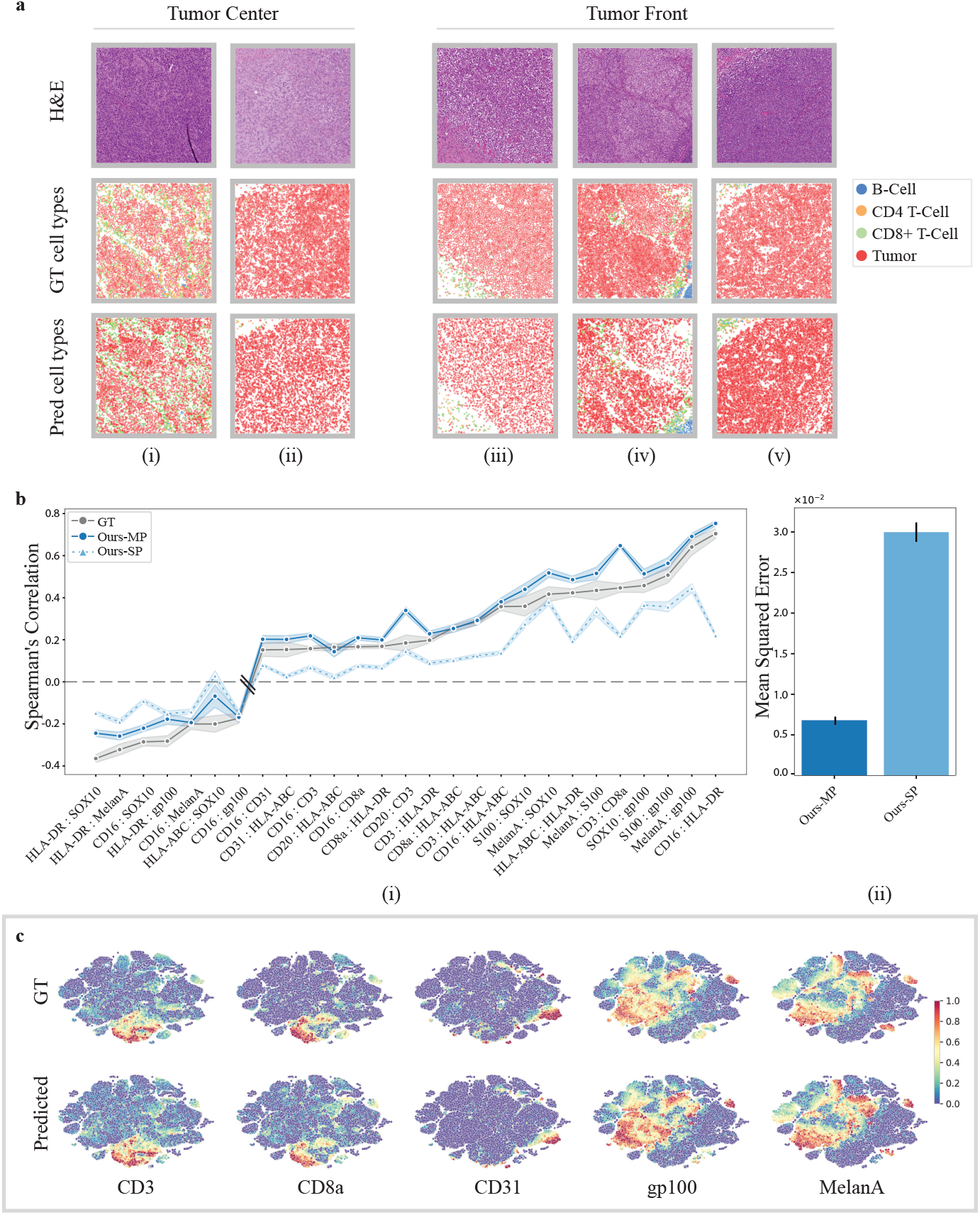
**a** Cell-typing results: H&E (first row), GT and predicted cell types (middle and bottom row) in RoIs grouped by their location within the tissue: “Tumor Center” and “Tumor Front”. **b**(i) Spearman’s correlation coefficients between protein pairs, comparing the ground truth (GT) with both singleplexed (SP) and multiplexed (MP) predictions of the HistoPlexer. The pairs on the X-axis are ordered by increasing Spearman’s correlation in the GT. **b**(ii) Mean squared error between the GT and predicted Spearman’s correlation coefficients, comparing the SP and MP predictions of the HistoPlexer. **c** Joint t-SNE visualization of protein co-localization patterns for selected markers: CD3, CD8a, CD31, gp100 and MelanA. The color represents protein expression.

### 2.3 HistoPlexer preserves spatial co-localization patterns

Given the importance of spatial patterns, as highlighted by [31, 32], we assess spatial co-localization by quantifying the correlations between protein markers that are simultaneously expressed within a given region. For each protein pair, we compute the Spearman’s Correlation Coefficient (SCC) between the two proteins and average it across RoIs, considering only those pairs with strong positive (*>* 0.15) or strong negative (*< —*0.15) correlation in GT IMC images. We then compare the SCC between GT and generated IMC multiplexes.

As shown in Fig. 3b(i), the Multiplex (MP) model’s predictions align more closely with the GT than those of the Singleplex (SP) model in terms of pairwise SCC, particularly for protein pairs involving CD-based immune markers such as CD16:HLA-DR, CD3:HLA-ABC and CD16:CD8a, which are underrepresented in the training data. We hypothesize that sparse markers lack sufficient tissue context for the SP model to generate accurate predictions. In contrast, the MP model benefits from learning inter-marker correlations by predicting all markers simultaneously. By leveraging auxiliary morphological information from abundant markers, it enhances predictions for both sparse markers and spatial co-localization. However, for some protein pairs (e.g., CD3:CD8a and CD20:CD3), the SCC in MP exceeds that of the GT. This is likely due to the similar morphological features of CD8+ T-cells (a subset of CD3 T-cells) and CD3 T-cells, as well as of B-cells (CD20) and CD3 T-cells in H&E images [28], potentially leading to overprediction of sparse markers and overestimated co-localization patterns. We further quantify spatial co-localization by measuring the Mean Square Error (MSE) between the SCC values from GT and generated IMC data across all test RoIs (Fig.3b(ii)). The MP model achieves an MSE approximately an order of magnitude lower than that of the SP model, supporting our hypothesis. Comparisons with Pix2Pix [23] and PyramidP2P [12] baselines is shown in Extended Data Fig. 2a. For the biological implication of observed patterns, refer S1.3.1.

To explore spatial patterns beyond protein pairs, we visualize expression profiles using t-SNE embeddings of cells from both GT and generated IMC multiplexes, following [33]. We observe a strong correspondence between t-SNE embeddings from both GT and generated IMC multiplexes (Fig.3c). For instance, cells that are positive for CD3 and CD8a are concurrently negative for CD31, gp100 and MelanA. This aligns with biological expectations, as CD3 and CD8a are expressed on T-cells but not on endothelial (CD31) or tumor (gp100, MelanA) cells. Full t-SNE plots for all markers are provided in Extended Data Fig. 2b.

In conclusion, our quantitative and qualitative results suggest that the spatial co-localization patterns in GT can be effectively replicated using the generated IMC images. These spatial patterns remain consistent across tissue sections, providing a robust evaluation metric that mitigates slice-to-slice variations.

### 2.4 HistoPlexer enables multiplexed proteomics profiling on the WSI-level

HistoPlexer enables the generation of IMC images from H&E WSIs of up to 100,000×100,000 pixels, allowing for the simultaneous visualization of multiple protein markers across entire tissue sections. This capability offers a comprehensive view of tumor and TME interactions at the WSI level. Since GT IMC data is available only for RoIs, we use Ultivue’s InSituPlex® technology to obtain multiplexed WSIs using the Immuno8 and MDSC FixVue™ panels. These panels include key markers, such as SOX10 for tumors, HLA-DR for antigen presentation, and CD3/CD8a for T-cell profiling, which are shared with the generated protein multiplex. Fig. 4 provides a qualitative comparison between the generated IMC and Ultivue multiplex at the WSI level. In both cases, a high concordance in global structures and hotspot regions is observed across all markers. In Fig. 4(ii), while there is good alignment for CD3 and SOX10 markers, differences are observed for CD8A and HLA-DR, particularly at the tissue edges (e.g., the bottom-left border). These differences are likely due to slice-to-slice variations between H&E and Ultivue images, which lead to slight shifts in tissue boundaries.

**Fig. 4:**
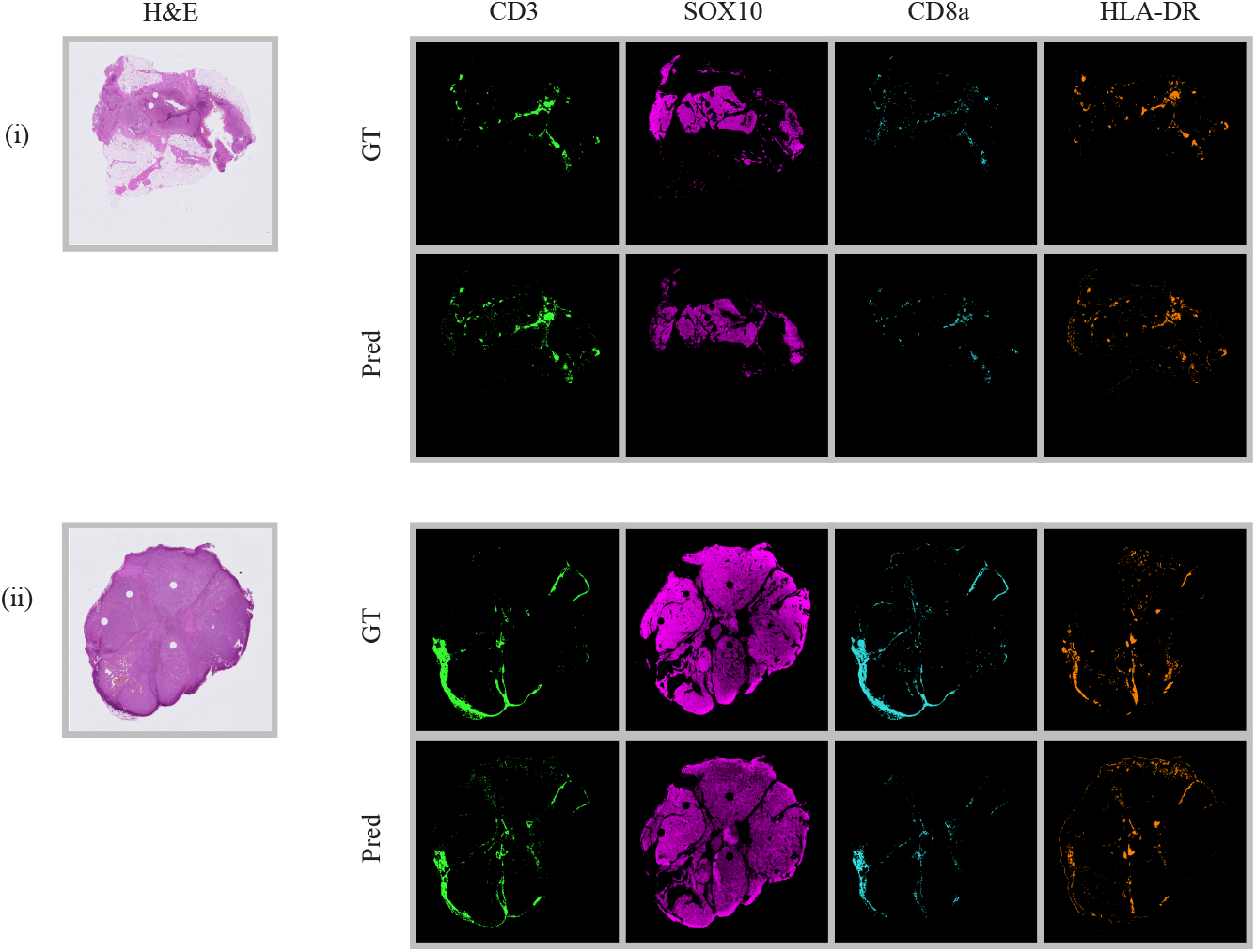
Qualitative WSI-level assessment of HistoPlexer. H&E (first column) and expression profiles of individual markers: CD3, SOX10, CD8a and HLA-DR (from second to last column). Top row: GT expression profiles from Ultivue images; bottom row: predicted (pred) expression profiles on WSI level both samples in (i) and (ii).

### 2.5 HistoPlexer facilitates immune phenotyping

We demonstrate the utility of HistoPlexer for immune phenotyping by stratifying immune subtypes based on the spatial distribution of CD8+ T-cells, using only H&E images from TuPro metastatic melanoma samples. As illustrated in Fig.5a, we visualize predicted tumor cells and CD8+ T-cells across H&E WSIs. In immune-hot cases, characterized by substantial CD8+ T-cell infiltration and typically associated with better response to immunotherapy [34, 35], we observe both tumor (attacker) cells and infiltrating CD8+ (defender) T-cells within the tumor region, suggesting an active immune response. In contrast, immune-cold cases exhibit minimal or no CD8+ T-cell infiltration in the tumor area, correlating with poor immunotherapy outcomes. Building upon the immune subtype classification framework proposed in [5], we further quantify intratumoral (iCD8) and stromal (sCD8) CD8+ T-cell densities within the tumor center compartment. To do this, we first localize CD8+ T-cells using HistoPlexer, then annotate and segment the tumor center into intratumoral and stromal regions using the HALO^AI^ platform across 34 TuPro melanoma samples.

Fig. 5b(i) shows immune subtype stratification using iCD8 and sCD8 densities (measured per *µm*^2^). We observe that immune desert cases exhibit very low iCD8 and sCD8 density, indicating the presence of only rare or isolated CD8+ T-cells. Immune excluded cases also show very low iCD8 density but slightly higher sCD8 density compared to immune desert cases, suggesting some CD8+ T-cells have reached the stroma but not the intratumoral regions. Inflamed cases display high densities of both iCD8 and sCD8, indicating the presence of CD8+ T-cells in the stromal compartment and, most importantly, their infiltration into intratumoral regions. These trends are consistent with prior findings [5], validating the utility of our approach. To assess the clinical relevance, we analyze the distribution of iCD8 and sCD8 densities across immune-hot (inflamed) versus immune-cold (excluded and desert) cases (Fig. 5b(ii)). As expected, immune-hot cases exhibit significantly higher densities of both iCD8 and sCD8 compared to immune-cold cases. Furthermore, a random forest classifier trained to distinguish between immune-hot and immune-cold cases achieves an F1 score of 0.873 (SD 0.006) and a macro-average AUROC of 0.845 (SD 0.047) under 5-fold cross-validation. In conclusion, we demonstrate that HistoPlexer can accurately support immune phenotyping directly from H&E images. This capability offers potential utility in treatment decision-making and patient stratification. For clinical implications of immune phenotyping, refer Section S1.3.2.

**Fig. 5:**
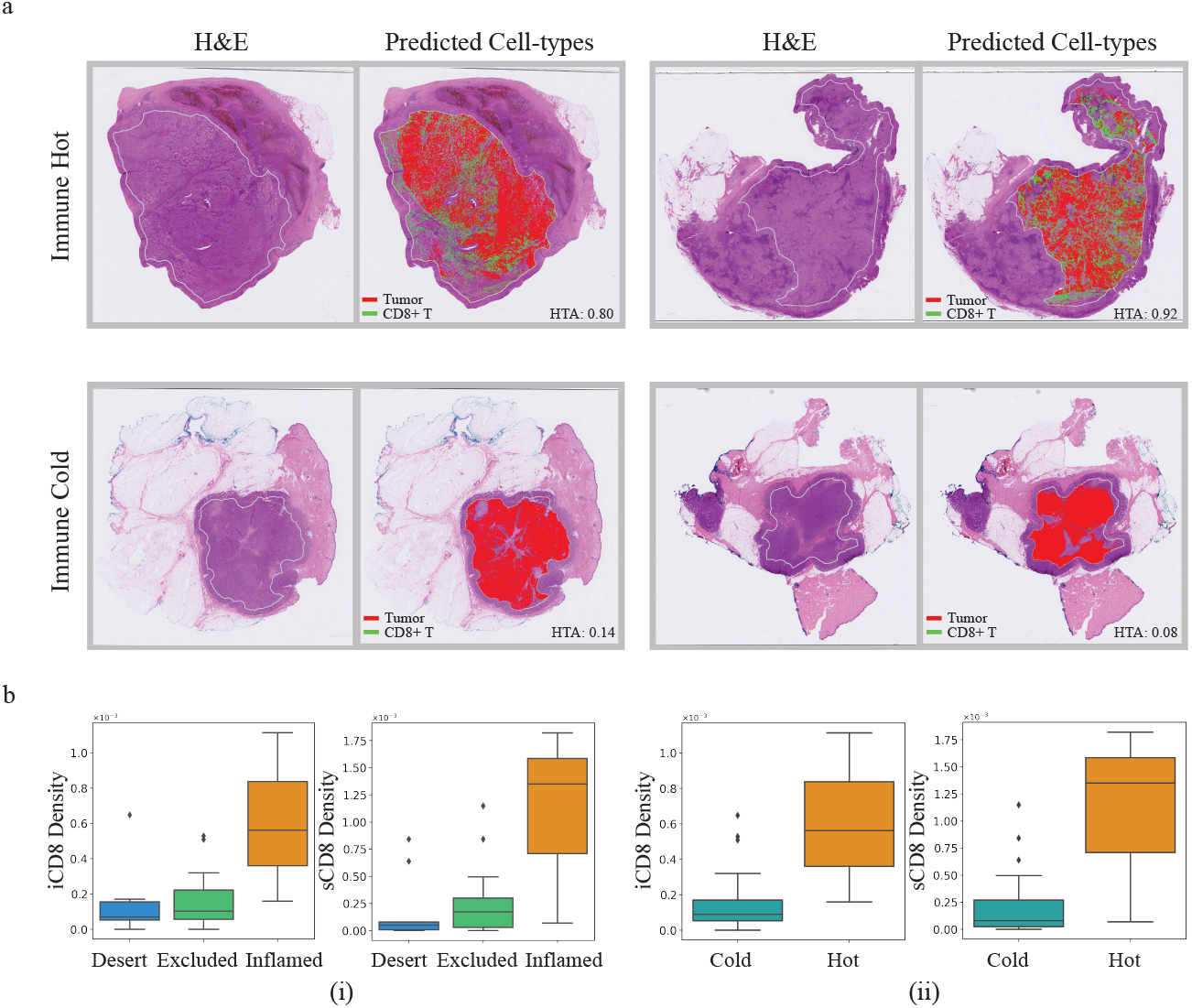
Immune phenotyping using HistoPlexer. **a** Four samples from TuPro metastatic melanoma cohort with two immune hot cases in the top row and two immune cold samples in the bottom row. For each sample, H&E image on left along with overlay of predicted tumor and CD8+ T-cells within tumor center region using HistoPlexer model on right. **b**(i) Box plot of intratumoral (iCD8) and stromal (sCD8) CD8+ T-cell densities in tumor center compartment, stratified by immune desert, excluded and inflamed classes. **b**(ii)Box plot of intratumoral (iCD8) and stromal (sCD8) CD8+ T-cell densities in tumor center compartment, stratified by immune hot and cold classes.

### 2.6 HistoPlexer generalizes to an independent patient cohort

We evaluate the generalizability of the HistoPlexer on independent test data from the TCGA-SKCM cohort [19]. Fig. 6a shows the generated protein multiplex at the WSI level, along with expression profiles for three markers: tumor-associated MelanA, T-cell marker CD3, and B-cell marker CD20. In the immune-high sample, we observe higher expression and tumor infiltration of CD3 and CD20 markers, contrasting with the minimal or absent expression in the immune-low case, where immune labels are based on Bulk RNA-seq data [36].

**Fig. 6:**
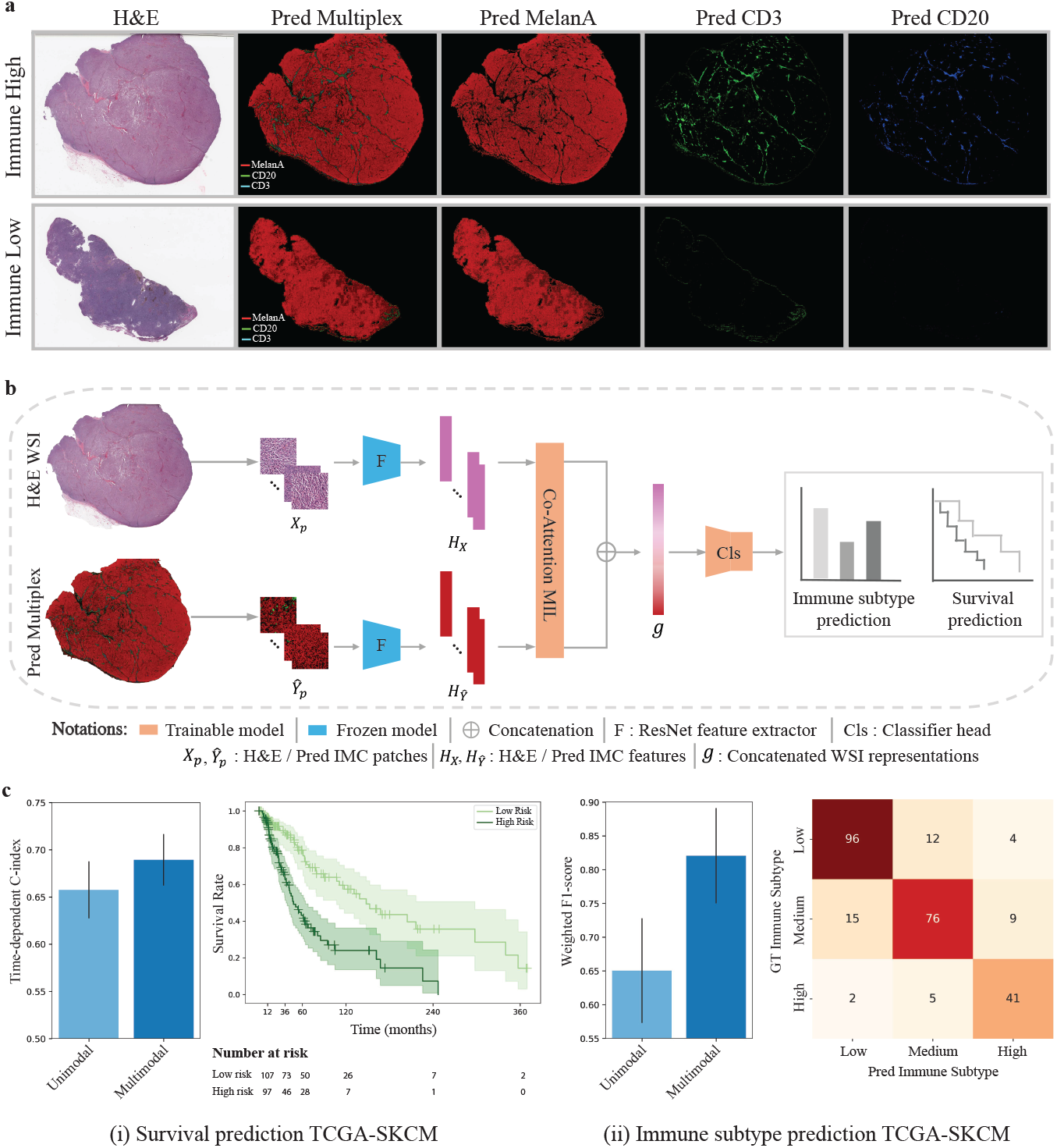
Generalization to an independent patient cohort. **a** Two examples (immune-high and -low) from the TCGA-SKCM cohort, showing H&E images (first column), predicted protein multiplexes (second row) as well as expression profiles of MelanA, CD3 and CD20 markers (last three colums). **b** Model architecture for multimodal survival and immune subtype prediction. **c(i)** Survival prediction results, displaying time-dependent c-index scores (left) and Kaplan-Meier survival curves for the multimodal setting, with separation of low- and high-risk groups (right).; **c(ii)** Immune subtype prediction results, showing the weighted F1 score (left) and confusion matrix (right) for classification into low, intermediate, and high immune subtypes.

To explore the clinical utility of the generated IMC images, we assess their value in augmenting outcome prediction tasks using the MelanA, CD3, and CD20 expression profiles, which are known to have prognostic significance [37, 38]. We extract features from both the H&E and generated IMC WSIs using pretrained encoders and input them to an attention-based Multiple Instance Learning (MIL) predictor [39]. We train the MIL predictor under two settings: (1) the unimodal setting, where only H&E features are input to the predictor and (2) the multimodal setting, where features extracted from the corresponding H&E and predicted IMC patches are first aggregated via a co-attention layer [40], and the bag-level representations of H&E and predicted IMC WSIs after the MIL pooling layer are concatenated before fed into the classification head (Fig. 6).

We benchmark the model on two clinically relevant tasks: immune subtype classification and survival prediction. For the survival task, we use disease-specific survival from patient metadata, which offers a more accurate representation of the patient’s disease status [41]. For immune subtyping, we classify patients into low, intermediate, and high immune categories based on Bulk RNA-seq-derived labels [36]. Overall, the multimodal setting consistently outperforms the unimodal configuration for both tasks. For survival prediction, incorporating features from generated IMC images yields a 3.18% improvement in average time-dependent C-index [42] across 5-fold cross-validation. Kaplan–Meier curves generated from the multimodal model show clear stratification of patients into low- and high-risk groups based on predicted risk scores (definition provided in 4.6), with statistical significance confirmed by a log-rank test (p-value = 5.05 × 10^−7^). For immune subtype classification, combining features from H&E and generated IMC boosts the average weighted F1 score by 17.02% over 5-fold cross-validation. In summary, these findings highlight both the generalizability of HistoPlexer to external cohorts and the clinical utility of the generated protein expression maps in supporting downstream predictive modeling and decision support.

## 3 Discussion

In this study, we introduced HistoPlexer, a generative model designed to predict high-order (11-channel) multiplexed protein expression profiles, including both tumor and immune markers, directly from H&E images. By predicting multiple proteins simultaneously, HistoPlexer effectively captured sparse markers and preserved biologically meaningful relationships, as validated through spatial correlation analysis of protein co-localization patterns. Our comprehensive evaluation demonstrated that the multiplexed prediction approach consistently outperformed singleplex alternatives, as reflected in higher MS-SSIM and PSNR scores and lower RMSE-SW and MSE values. Notably, domain experts found the generated IMC images realistic, with HYPE error rates of 61.6% and 72.8% for lymphocyte and tumor markers, respectively, supporting the quality of our predictions. To assess the broader applicability of HistoPlexer across different cancer types, we benchmarked it on the DeepLIIF datasets [14, 20], which provide pixel-aligned H&E and multiplexed images for lung and head and neck cancers. In both settings, HistoPlexer consistently outperformed baseline methods, indicating robustness across diverse tissue types and imaging conditions (Extended Data Tables 1 and 2).

We further explored the integration of features from the UNI computational pathology foundation model (FM) [43] to enhance the performance of HistoPlexer, marking one of the first attempts to apply pathology FMs to generative tasks. As outlined in Section S1.2.1 and shown in Extended Data Table 3, end-to-end training of HistoPlexer without UNI-derived features resulted in lower MSE and better preserved biologically relevant relationships. This finding are consistent with recent studies [44] and underscores the importance of task-specific training for complex biological applications. Foundation models, typically designed as encoder-only architectures, excel in discriminative tasks like classification and regression by focusing on global high-level features. However, they are less suited for generative tasks that require detailed spatial relationships and fine-grained image details essential for accurate image generation.

We demonstrated the clinical utility of HistoPlexer through two key applications. First, HistoPlexer enabled immune phenotyping at the WSI level by quantifying spatial patterns of CD8+ T-cell infiltration using intratumoral (iCD8) and stromal (sCD8) densities in the tumor center compartment. The spatial patterns were in concordance with the state-of-the-art approach [5], and were effective in stratifying patients into clinically actionable immune hot and cold subtypes. This capability is particularly valuable for immunotherapy decisions, where understanding the spatial distribution of CD8+ T-cells is crucial. Second, we showed generalizability to an independent cohort (TCGA-SKCM) where HistoPlexer-generated features, when used alongside H&E features, improved predictive performance in survival analysis (3.18% increase in time-dependent C-index) and immune subtype classification (17.02% improvement in weighted F1 score). These results highlight the potential of HistoPlexer to augment clinical decision-making.

This study has some limitations. First, in some cases, the model confused morphologically similar markers, such as CD3/CD8a T-cells and CD20 B-cells. While this did not significantly impact downstream tasks, it is a priority for future work to refine the model’s ability to accurately distinguish between finer cell types. Second, our focus was on major immune and tumor cell types, such as CD8+ T-cells, CD4+ T-cells, B-cells, and tumor cells. This set could be further extended to include more sparse cell types, such as endothelial cells by obtaining a larger training cohort. Third, for the multimodal learning experiments on TCGA-SKCM, we limited marker selection to MelanA, CD3, and CD20 based on clinical relevance. Extending the marker panel could further improve downstream applications. Lastly, due to inherent slice-to-slice variations, we prioritized downstream task performance over strict pixel-level accuracy.

Our work opens several promising research directions. First, expanding HistoPlexer to additional protein markers and cancer types presents an opportunity to uncover deeper insights into disease mechanisms without requiring additional tissue or increased cost. Achieving broader generalization, including the development of a universal model across cancer types, remains a key challenge that will require access to larger, more diverse, and harmonized datasets. Our current results serve as a proof of concept and provide a foundation for such future efforts. Second, by releasing the Ultivue InSituPlex® dataset generated in this study, we encourage researchers to explore novel diffusion models for multiplexed protein marker generation, particularly those that account for slice-to-slice variations. Finally, integrating predicted protein expression maps with other molecular modalities holds the potential to deepen our understanding of tumor biology and improve patient stratification.

In conclusion, HistoPlexer represents a significant advance in computational pathology, enabling cost-effective generation of protein multiplexes from clinically established histology slides. Our results support its potential for real-world clinical integration, with significant implications for improving cancer diagnosis and guiding personalized therapy.

## 4 Methods

### 4.1 Datasets and preprocessing

#### 4.1.1 Tumor Profiler dataset

We build our HistoPlexer framework using a subset of highly multi-modal metastatic melanoma dataset generated by the Tumor Profiler Study (TuPro) [18]. Each patient was characterised using multiple technologies, including Digital Pathology and IMC. A total of six RoIs of 1 mm^2^ were selected on each H&E WSI, three within tumor center and three at the tumor front (intersection of tumor and TME). IMC data was generated for those six RoIs on a consecutive section of the same tumor block. The IMC data was generated at a resolution of 1µm/pixel and H&E images were scanned at a resolution of 0.25 µm/pixel. Therefore, RoIs of 1 mm^2^ are represented by 1000 pixels for IMC data and 4000 pixels for H&E images. Since the paired data was generated by visually choosing RoIs, in many cases a considerable positional shift and rotation between the specified H&E regions and the resulting IMC regions can be observed. This was overcome by using template matching [45]. Specifically, for the IMC modality, we utilized the tumor channel marker SOX10, which provided valuable structural information of the tissue. For the H&E modality, we focused on the H channel, which captures the hematoxylin staining essential for visualizing cellular structures. Given that H&E and IMC represent different modalities, we employed mutual information as a metric for template matching. This approach allowed us to quantify the alignment between the two datasets, ensuring a robust pairing process. This resulted in a paired dataset of 336 H&E and IMC ROIs from 78 patients for training and testing model performance.

IMC profiling was performed using a panel of 40 antibodies, from which 11 have been selected for this study based on the biological function of the corresponding proteins as well as high signal–to–noise ratio. The proteins targeted by the 11 antibodies include cell-type markers, such as tumor markers (MelanA, gp100, S100, SOX10), lymphocyte markers (CD20, CD16, CD3, CD8a) and an endothelial marker (CD31). Moreover, two functional markers corresponding to proteins involved in antigen presentation (HLA-ABC, HLA-DR) are included in the protein set.

The raw IMC images were processed with CellProfiler software for cell segmentation [46]. The protein counts extracted from the images have been first clipped to 99.9% per protein to exclude outliers ad then transformed using the *arcsinh*-function with cofactor one [47]. In order to exclude background noise, we apply OTSU thresholding [48] with kernel size three and sigma three and the threshold, separating signal from background, determined per sample using all available RoIs. The resulting data per protein is first centered and standardized and then subjected to min-max-transformation, all using data statistics based on the train set only.

The data is split at the patient level into train and test sets, stratified by immune phenotype (inflamed, immune excluded, and immune desert). The stratification ensures the representation of both tumor and immune cells in each set. The patient-level splitting guarantees that all RoIs from a given patient belong to only one set, preventing undesired information flow. The resulting train and test sets consist of 231 and 105 RoIs, respectively. During model training, RoIs are chosen at random and a tile of size 1024×1024 from H&E image and a corresponding IMC region of 256×256 is extracted.

For whole-slide image (WSI) inference, we developed a modular pipeline that processes WSIs in a tile-based approach to create spatially-resolved protein multiplexes. The process begins with tissue segmentation, which identifies stained tissue areas and removes the background using OTSU thresholding [48]. Each segmented tissue region is then divided into 1024×1024 pixel tiles. To optimize performance, we employ a dataloader that batches and parallelizes the processing of these tiles instead of processing them sequentially. We also utilize the Shapely library [49] for geometric operations, such as creating polygons for tissue masks, scaling them to match the WSI resolution, and efficiently determining overlaps between tiles and tissue regions. This approach reduces computational load by concentrating only on relevant areas of the WSI and avoiding background processing. Each tile may undergo optional stain normalization using the Macenko method [50] to reduce staining variability and ensure color consistency across images. The tiles are then processed through the trained HistoPlexer model to predict protein multiplexes. After prediction, the generated tiles are stitched together to reconstruct the full WSI at multiple resolutions. We also generate quality control images to visualize the tiling and segmentation results, and we save the coordinates of the tiles for reproducibility. For visualization, we apply min-max normalization using the min-max values from the training set for all markers.

#### 4.1.2 Ultivue dataset

For qualitative evaluation of HistoPlexer on WSIs, we employed Ultivue InSituPlex® technology to obtain multiplexed images using the Immuno8 and MDSC FixVue™ panels. The Immuno8 panel focuses on immune landscape characterization with markers such as CD3, CD4, CD8, CD68, PD-1, PD-L1, FoxP3, and PanCK/SOX10. The MDSC panel identifies myeloid-derived suppressor cells using markers CD11b, CD14, CD15, and HLA-DR. Ultivue images were acquired at a resolution of 0.325 µm/pixel. For evaluation, we used CD3, SOX10, CD8a, and HLA-DR markers to assess visual similarity between the generated protein multiplex and Ultivue images.

Paired H&E and Ultivue WSIs were generated by first staining H&E on one tissue section, followed by acquiring Immuno8 and MDSC data on consecutive sections for 10 samples. A tonsil tissue was included with each sample as a positive control. Image registration between H&E and Ultivue WSIs was performed using an unsupervised multimodal method [51], leveraging the DAPI nuclear stain in Ultivue for alignment with H&E images. Both Ultivue and generated IMC images underwent minmax normalization and histogram equalization. Additionally, adaptive thresholding was applied to Ultivue images to reduce noise and extract true signal. Regions with false signals, particularly those corresponding to hemorrhage, bleeding, or erythrocytes in H&E, were manually annotated and excluded from the analysis. Upon acceptance, we plan to publicly release the H&E and Ultivue images, their alignment matrices, and annotated excluded regions. The dataset could serve as a valuable baseline for the field.

#### 4.1.3 TCGA-SKCM

Diagnostic WSIs of SKCM were downloaded from the TCGA database^1^ for a total of 472 cases. Clinical data of SKCM samples including age, gender, sample type (primary tumor/metastatic) and disease-specific survival were also downloaded. For the survival prediction, we discarded cases where the diagnostic WSIs are of low resolution or the disease-specific survival data is missing, leaving 360 cases in total. For the immune subtype prediction, we kept a total of 257 cases where immune subtype labels are available. For each task, we randomly split the cases stratified by age, gender and sample type to create 5-fold cross-validation with a 4:1 ratio of training-validation sets.

### 4.2 HistoPlexer architecture

The HistoPlexer is based on cGAN which takes an H&E image as input condition and generates multiplexed IMC images where each corresponds to a spatially-resolved protein expression profile. The *translator* of the HistoPlexer is a fully convolutional U-Net [52] which consists of an encoder and a decoder. The encoder comprises six downsampling blocks, each with a convolution layer of stride 2 and kernel size 3. The decoder comprises of five upsampling blocks, each with nearest neighbor interpolation, followed by convolution layer of stride 1 and kernel size 3. Each layer is followed by a batch-norm layer and ReLU activation. The *discriminator* consists of six blocks, each with a convolution layer followed by a spectral normalization layer and ReLU activation. We use patches extracted from template-matched pairs of H&E and IMC RoIs to train the HistoPlexer and optimize the model with three objectives: an adversarial loss to enforce image-level consistency, a Gaussian pyramid loss to enforce pixel-level consistency, and a patch-wise contrastive loss to enforce patch-level consistency.

#### Adversarial loss

We use the least square loss proposed in LSGAN [53] as our adversarial loss, and the 0*—*1 coding scheme where 0 and 1 are the labels for generated (*i.e*., fake) and real IMC images, respectively. We also adopt the multi-scale gradient approach [54], which allows simultaneous gradient propagation at multiple scales (*i.e*., resolutions). Considering a set of scales {*s* ∈ *S*}, the multi-scale adversarial losses for the translator *G* and discriminator *D* are formulated as:

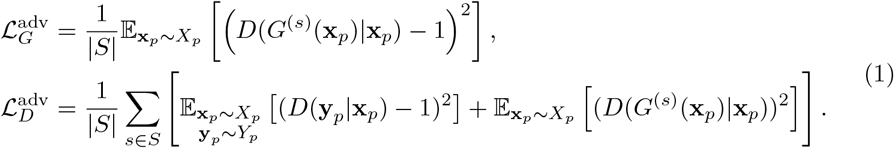

where *X*_*p*_ = {**x**_*p*_ ∈ *X*_RoI_} and *Y*_*p*_ = {**y**_*p*_ ∈ *Y*_RoI_} denote paired training patches sampled from template-matched H&E and IMC RoIs, respectively; *G*^(*s*)^(·) and *D*(·) denote the mapping functions parameterized by the translator (at the output scale *s*) and discriminator, respectively; and | · | denotes the cardinality of a set.

#### Gaussian pyramid loss

We also implement a pixel-level *L*_1_ loss as in [23]. Since our H&E and GT IMC images are not pixel-aligned, we relax the constraint on pixel-to-pixel correspondence by calculating the *L*_1_ loss at multi-resolution representations of the generated and GT IMC images [12], termed as Gaussian pyramid loss [12]. More specifically, a Gaussian pyramid is constructed through iterative Gaussian smoothing and downsampling. Each level of resolution, termed as an octave, comprises a series of images with increasing degrees of smoothness. Transition between resolutions is achieved by downsampling the image at the highest smoothness level of the current octave to initiate the next:

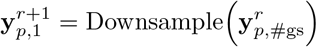

where #*gs* denotes the number of Gaussian smoothing at one resolution. Note that for the generated IMC images, we only compute the Gaussian pyramid on the final output scale. Considering a set of resolutions {*r* ∈ *R*}, the Gaussian pyramid loss is a weighted sum of *L*_1_ loss computed on the primary layer of each octave, formulated as:

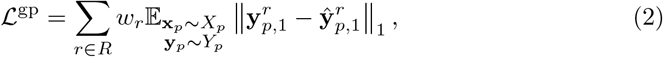

where **ŷ**_*p*_ denotes the generated IMC image patches, *r* denotes the resolution level, and *w*_*r*_ is the weight of the *L*_1_ loss at that level.

#### Patch-wise contrastive loss

We further incorporate a patch-wise contrastive loss, inspired by [21]. More specifically, we first extract multi-layer features using a pretrained feature encoder and apply a transformation via a small projection head (*e.g*., a Multi-layer Perceptron) on the extracted features to enrich their expressiveness [55]. Then, we randomly select a set of pixel locations for each feature layer. By aggregating selected patch features from each layer, we can obtain two feature sets for the generated and GT IMC images, respectively.

Let 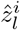 denote the anchor feature of the *i*-th patch of the generated IMC image, extracted from the *l*-th layer of the feature encoder; while 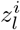 and 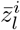 denote the positive and negative features of the corresponding patch (*i.e*., at the same pixel location) and the collection of non-corresponding patches (*i.e*., at different pixel locations), extracted from the same layer, respectively. Our patch-wise contrastive loss is defined as:

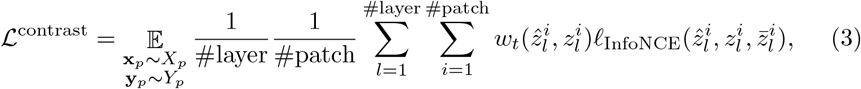

where

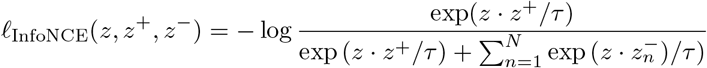

is the InfoNCE objective [56], and

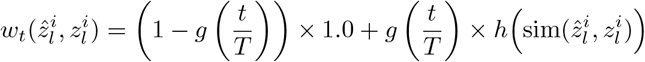

is the adaptive patch weight [21]. Here, #layer and #patch denote the number of layers and patches from which we extract features; *t* and *T* denote the current and total training steps; *h*(·) denotes some weighting function; and sim(·) is some similarity measurement.

While the HistoPlexer translator outputs the prediction of all selected IMC markers, we encounter a practical limitation when employing a pre-trained feature encoder, which often requires an RGB image as input. To circumvent this, we first extract each channel (*i.e*., marker) of the output IMC image and replicate it along the channel dimension to create a pseudo RGB image. We then pass each of them to the feature encoder. The final patch-wise contrastive loss is the sum of that of each channel.

The total losses for *G* and *D* are formulated as,

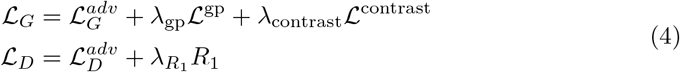

where

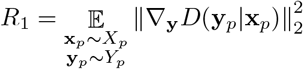

is the gradient penalty [57], and *λ*_gp_, contrast and 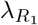 are the weights for the Gaussian pyramid loss, patch-wise contrastive loss and gradient penalty, respectively.

#### Implementation and training details

The model is trained for 100 epochs using ADAM optimizer [58] with momentum parameters *β*1 = 0.5 and *β*2 = 0.999 with learning rates 0.004 and 0.0008 for translator and discriminator networks, respectively. The weights are initialized using Xavier initialization. The batch size is set to 16 and the patch size to 256 for IMC and 1024 for H&E images, to accommodate for the higher resolution of the latter. We increase the generalization capabilities of the model by adopting data augmentation, including color augmentation, random flipping, small translations, and rotations. We employ the least-squares GAN objective. The weights for loss terms is as follows: *λ*_gp_=5.0, *λ*_contrast_=1.0 and 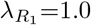. For model inference and evaluation, we use the latest checkpoint at the end of model training.

### 4.3 Evaluation metrics

To evaluate the quality of generated images, we use three widely adopted metrics: PSNR, MS-SSIM and RMSE-SW.

PSNR is used to measure the reconstruction quality by quantifying the ratio between the maximum possible signal power and the power of corrupting noise. It is expressed in decibels (dB), with higher values indicating better image quality. The PSNR is calculated as:

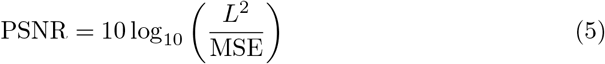

where *L* is the dynamic range of the pixel values (e.g., 255 for 8-bit images), and MSE represents the Mean Squared Error between the original image *I* and the generated image *I*′

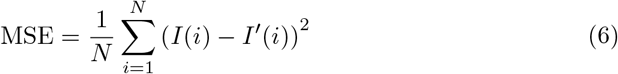

MS-SSIM extends the traditional SSIM metric by incorporating multiple scales to capture structural differences at various resolutions. The SSIM between two images *I* and *I*′ is defined as:

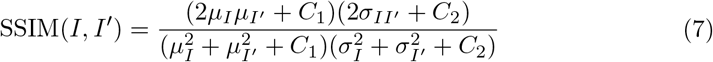

where *µ*_*I*_ and 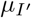 are the means, 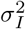 and 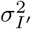 are the variances, and *σ*_*II*′_ is the covariance between the two images. *C*_1_ and *C*_2_ are small constants to stabilize the division. In MS-SSIM, is computed at multiple scales, and the final score is a weighted product of SSIM values across these scales:

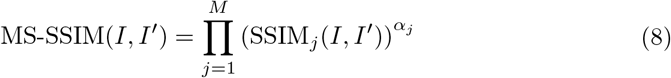

where *M* is the number of scales and *α*_*j*_ is weighting factor at scale *j*. Higher MS-SSIM values indicate better perceptual similarity.

RMSE-SW is an image quality metric used to assess the similarity between the original image *I* and a generated image *I*′. Unlike traditional RMSE, which computes the error over the entire image globally, RMSE-SW calculates the error within localized regions by moving a fixed-size window across the image. The RMSE-SW between two images *I* and *I*′ is defined as:

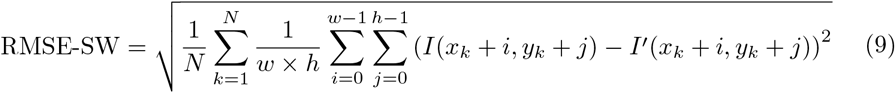

where w × h represents the size of the moving window across the image with a defined stride, resulting in *N* windows. For each window, we compute the mean squared error between the corresponding pixel values of *I* and *I*′.

These metrics provide a comprehensive assessment of both pixel-level accuracy and perceptual similarity of the generated images. Frechet Inception Distance (FID) and Kernel Inception Distance (KID) are widely used metrics for evaluating the quality of generated images, however, they are less effective on small datasets as they rely on mean and covariance of a cohort. Hence they are not used when evaluating HistoPlexer.

To quantify the evaluation by domain experts, we use HYPE score which measures the error rate at which humans mistake generated images for real ones or vice versa. It is defined as:

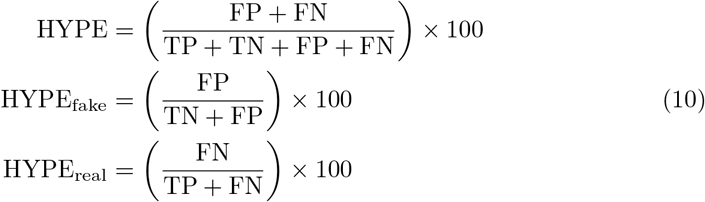

where TP is the number of True Positives, TN is the number of True Negatives, FP is the number of False Positives and FN is the number of False Negatives. HYPE_*fake*_ and HYPE_*real*_ are the error rates for generated and real images, respectively.

### 4.4 HistoPlexer for cell-level analysis

#### 4.4.1 Pseudo-cells

Since spatial analyses of IMC data typically rely on cell-level readouts, we create pseudo-single-cell data by extracting circular regions of 10 µm diameter around nuclei coordinates for both input H&E and GT IMC images. Protein expression is averaged across pixels within each pseudo-cell for individual markers. Nuclei coordinates for H&E images are obtained using the HoVer-Net model [28], while nuclei coordinates and cell-type labels for GT IMC multiplexes are derived using Ilastik [59] and Cell-Profiler [46], as described in [8]. For simplicity, we refer to pseudo-cells as “cells” in the following text.

#### 4.4.2 Cell-typing

We use a Random Forest (RF) classifier [30] to categorize cells based on the average expression of 11 markers from the HistoPlexer. The classifier distinguishes between tumor cells, B-cells, CD8+ T-cells, CD4+ T-cells, and other cells. Training is performed using the scikit-learn library [60], with hyperparameters (100 base estimators, maximum tree depth of 30) selected based on the lowest out-of-bag error. The model achieves a macro-averaged F1 score of 0.81 on an internal test set. We then apply the trained RF classifier to both GT and generated protein expression data to produce cell type maps for cells in test set.

#### 4.4.3 t-SNE on cell level marker expression

To explore spatial patterns beyond pairwise protein interactions, we conduct a low-dimensional embedding analysis of cell-level marker expression. Following the approach commonly used or mass cytometry data [61], we subsample 1,000 cells per RoI from both GT and generated IMC, resulting in total 2,000 cells per RoI. A joint t-SNE dimensionality reduction (two dimensions, perplexity of 50, and 1,000 iterations) is then applied. For visualization, protein abundance is scaled and clipped at the 99th percentile, and the t-SNE plots are colored according to the scaled protein expression [61].

### 4.5 Annotations for Immune phenotyping

To stratify samples into immune subtypes based on the spatial distribution of CD8+ T-cells, we used annotated regions as established in [5]. Our dataset included 109 metastatic melanoma H&E WSIs from the TuPro cohort, with metastatic sites in lymph nodes, soft tissue, brain, and other distant locations. The primary region for immune-subtyping, termed “Tumor Center”, comprises entirely tumor tissue, which was manually defined as a continuous tumor mass excluding a 500*µ*m margin from the tumor–non-tumor boundary. This “Tumor Center” was further segmented into two regions: the “Intratumoral Tumor” region, consisting of dense clusters of malignant melanocytes without stromal presence, and the “Intratumoral Stromal” region, which includes extracellular matrix (typically desmoplastic) interwoven within the tumor cell mass but free from malignant melanocytes. These regions were automatically classified using a DL model implemented on the HALO^AI^ platform, trained with selected H&E WSIs regions. Tissue classification was conducted at 0.30*µ*m/pixel resolution with a minimum object size threshold of 50*µm*^2^. Excluded regions—such as preexisting lymphatic tissue, large adipose and muscle regions, artifacts, necrosis, hemorrhage, and background—were omitted from the analysis. Ultimately, we analyzed 34 samples with the highest quality tissue classifications from the HALO^AI^ model predictions. Extended Data Fig. 3 shows an example H&E WSI with region annotation and classification.

### 4.6 MIL-based Clinical Outcome Prediction

#### Attention-based MIL for survival and immune subtype prediction

MIL is a weakly-supervised learning method for set-based data structures. In MIL, an input *X* is a bag (*i.e*., permutation-invariant set) of instances *X* = {**x**_1_, …, **x**_*N*_}, where *N* denotes the number of instances in the bag. Given a classification task with *K* classes, the goal is to learn a function *ℱ* from *M* training pairs 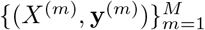 that maps *X* to a bag-level label **y** ∈ *K* without knowing label **y**_*i*_ ∈ *K* for each instance in the bag. In our context, the input is a WSI and the instances denote the extracted patches. More specifically, we follow the embedding-based MIL approach [39] and extract a feature vector **h**_*i*_ = *h*(**x**_*i*_) ∈ *ℝ*^*d*^ from each patch. Then, an attention-pooling operator aggregates the patch features **h**_*i*=1:*N*_ to a single WSI-level representation [39]

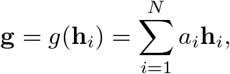

where

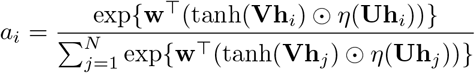

is the gated attention [39]. Here, **w**∈ℝ^*L×*1^, **V**∈ℝ^*L×D*^, **U**∈ℝ^*L×D*^ are learnable parameters with hidden dimension *L*, ⊙ is element-wise multiplication, and η (·) denotes the Sigmoid function. Finally, a classifier *f*(·) maps the WSI-level representation to a WSI-level label **Ŷ** ∈ *K*.

The end-to-end prediction takes the following general form:

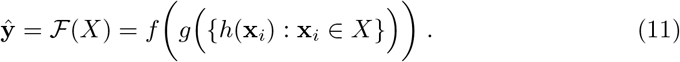

For survival prediction, we model the time-to-event distributions as an ordinal regression task with right censored data (*i.e*., patient death is unobserved until last known follow-up). Following [40], we define discrete time intervals and model each interval using an independent neuron in the output layer. More specifically, we partition the continuous time scale into non-overlapping time intervals [*t*_*j—*1_, *t*_*j*_),*j* ∈ [1, *…, J*] based on the quartiles of survival time values, denoted as **y**_*j*_. The continuous time-to-event *t*^(*m*)^ for each patient is then replaced by a discrete time label 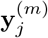, where

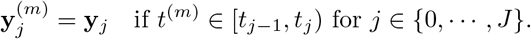

The problem then simplifies to classification where each patient is defined by a triplet (**g**^(*m*)^, **y**^(*m*)^, *c*^(*m*)^). Here, **g** is the aggregated bag features; *c* is the censorship status where *c* = 0 if the death of the patient is observed and *c* = 1 otherwise; and **y**_*j*_ is the discrete time GT label. We adopt the negative log-likelihood survival loss [62] for modal optimization, formulated as:

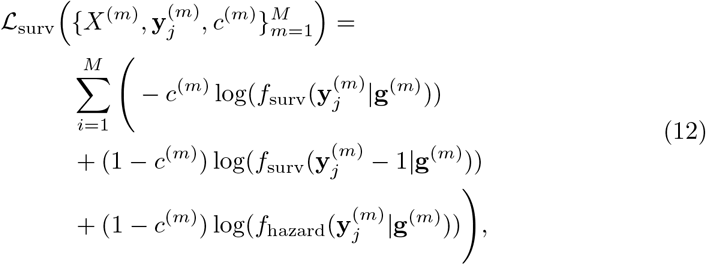

where *f*_harzard_(**y**_*j*_|**g**) = Sigmoid(**ŷ**_*j*_) is the discrete hazard function and 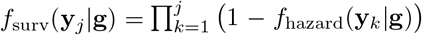 is the discrete survival function. Finally, the patient-level risk is defined as the negative sum of all logits [41], which enables the identification of distinct risk groups and the stratification of patients.

For immune subtype prediction, we adopt the cross-entropy loss defined as:

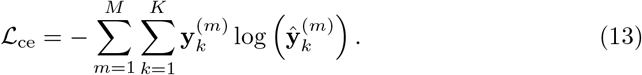

#### Multimodal fusion via co-attention mechanism

To fuse the patch features from different modalities, we adopt the co-attention mechanism proposed in [40]. More specifically, given the H&E feature bag **H** ∈ *ℝ*^*N×d*^ and IMC feature bag **P** ∈ *ℝ*^*N×d*^, we guide the feature aggregation of **H** using **P** by calculating the cross-attention:

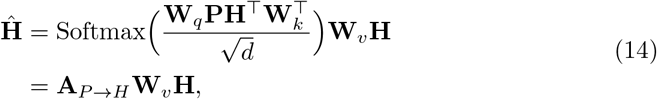

where **W**_*q*_, **W**_*k*_, **W**_*v*_ ∈ *ℝ*^*d×d*^ are learnable weights and **A**_*P→H*_ ∈ *ℝ*^*N×N*^ is the co-attention matrix. Intuitively, the co-attention measures the pairwise similarity for how much an H&E instance **h**_*i*_ attend to the IMC instance **p**_*i*_ for *i* ∈ *N*. Similarly, we can guide the feature aggregation of **P** using **H** via **A**_*H→P*_. Each co-attention guided feature bag is input to an attention-based MIL module, which outputs an aggregated WSI-level representation. We concatenate the WSI-level representations from multiple modalities and project it back to the original feature dimension *d* via a linear layer, resulting in a multimodal WSI-level representation. Then, a classifier *f*(·) uses this representation to predict the output label **ŷ**.

#### Implementation and training details

We adopt the original implementation of attention-based MIL on GitHub^2^ and modify it for survival prediction based on the code for SurvPath^3^. We implement the co-attention mechanism based on the original implementation of MCAT^4^. Each WSI is cropped to 256×256 non-overlapping patches at 20× magnification to create bags, where patches with more than 10% non-tissue area are discarded. We use ResNet18 [63] pretrained on pathology-specific datasets using self-supervised learning [64] to extract features from H&E patches and ResNet50 pretrained on ImageNet [65] to extract features from IMC patches. Since ResNet18 requires three-channel input, we concatenate IMC images of three different protein markers along the channel dimension: one tumor marker (MelanA) and two immune markers (CD8 and CD20). The dimension of extracted features is 512 for both H&E and IMC patches. We run the survival and immune subtype prediction for 5-fold cross-validation. The model hyperparameters are set as: Adam optimizer with initial learning rate of 1*e*^−4^ (survival) and 5*e*^−5^ (immune subtype), a ReduceLROnPlateau scheme based on validation loss for scheduling, and a mini-batch size of 1. The model is trained for 100 epochs with early stopping based on validation loss (survival) and weighted F1-score (immune subtype).

## Data Availability

Data and material from the TumorProfiler study are available to members of the Tumor Profiler Research Consortium. Requests for sharing of all data and material should be addressed to the corresponding author and include a scientific proposal. Depending on the specific research proposal, the Tumor Profiler consortium will determine when, for how long, for which specific purposes, and under which conditions the requested data can be made available, subject to ethical consent.

https://tumorprofilercenter.ch/

## Computational requirements

The data processing and model training was done on NVIDIA A100 40GB GPU. The DL models were trained using pytorch (1.13.1). The pipeline was implemented in Python (3.8.12).

## Data Availability

The Tumor Profiler study data used for training in the study is not publicly available. It is available to members of the international Tumor Profiler Research Consortium. Requests for sharing of all data and material should be addressed to the corresponding author (G.R.) and include a scientific proposal. Depending on the specific research proposal, the TumorProfiler consortium will determine when, for how long, for which specific purposes, and under which conditions the requested data can be made available, subject to ethical consent. The multiplexed WSIs images for Immuno8 and MDSC FixVue™ panels from Ultivue InSituPlex® technology, along with paired H&E images will be made available upon acceptance of publication. The H&E WSIs for TCGA-SKCM were downloaded via GDC data portal (https://portal.gdc.cancer.gov/).

## Code Availability

The source code for HistoPlexer is available at https://github.com/ratschlab/HistoPlexer and via Zenodo at https://doi.org/10.5281/zenodo.15110117 [66].

## Acknowledgements

We gratefully acknowledge funding from the Tumor Profiler Initiative and the Tumor Profiler Center (to V.H.K., G.R., B.B.). The Tumor Profiler study is jointly funded by a public-private partnership involving F. Hoffmann-La Roche Ltd., ETH Zurich, University of Zurich, University Hospital Zurich, and University Hospital Basel. We also acknowledge funding from the Swiss Federal Institutes of Technology strategic focus area of personalized health and related technologies project 2021-367 (to G.R., V.H.K., S.A.), the ETH AI Center (to G.R. and B.C.), ETH core funding (to G.R.), UZH, USB and UniBas core funding (to V.H.K) and funding by the Promedica Foundation grant F-87701-41-01 (to V.H.K). B.B. was funded by two SNSF project grants (#310030 205007: Analysis of breast tumor ecosystem properties for precision medicine approaches and #316030 213512: Cellular-resolution high-performance mass spectrometric imaging of biological samples), an NIH grant (UC4 DK108132), the CRUK IMAXT Grand Challenge, and the European Research Council (ERC) under the European Union’s Horizon 2020 Program under the ERC grant agreement no. 866074 (“Precision Motifs”).

We thank Marta Nowak and Anja Frei for their contributions to the generation and review of digital pathology data for immune profiling. We are also grateful to Susanne Dettwiler, Fabiola Prutek, Norbert Wey, and Sven Krackow from the Department of Pathology and Molecular Pathology, University Hospital Zurich, for their excellent collaboration and technical assistance. We acknowledge Ilario Scapozza (University of Zurich, Switzerland) for his support in the visual assessment evaluation, and Flavia Pedrocchi for her work on pathology image registration. We express our gratitude towards Lucy Godson (University of Leeds, UK) and Jeremie Nsengimana (Newcastle University, UK) for sharing the immune subtype data for TCGA-SKCM.

## Author Contributions

G.R., V.H.K, S.A. and J.F.P. conceived the study. G.R., V.H.K, S.A., B.C. and J.F.P. participated in designing the study and reviewing the literature. V.H.K and G.R. supervised the study. V.H.K., S.A., J.F.P., B.S. and R.C. collected the data. S.A., J.F.P., B.C. and S.H. contributed to code writing, model training, and data analysis. V.H.K. and G.R. contributed to expert review and data interpretation. B.C., S.A., V.H.K., and G.R. designed the figures. S.A. and B.C. drafted the manuscript. T.P.C., V.H.K, R.C., B.B. and B.S. contributed to providing data and tumor material used in this study. B.H. contributed towards annotation tasks and qualitative assessment of data. All authors had access to the result data presented in the final manuscript and all authors read and approved the final manuscript. The decision to submit was made by all authors. V.H.K. and G.R. contributed equally to this work and share senior authorship.

## Approval from ethics committee

The ethics committee of the “Swiss Association of Research Ethics Committees” gave ethical approval for the data from the Tumor Profiler Study used in this work. The Tumor Profiler Study is an approved, observational clinical study (BASEC: 2018-02050, 2018-02052, 2019-01326, 2024-01428).

## Distribution/reuse options

Anyone can share this material, provided it remains unaltered in any way, this is not done for commercial purposes, and the original authors are credited and cited.

## TUMOR PROFILER CONSORTIUM

Rudolf Aebersold^5^, Melike Ak^33^, Faisal S Al-Quaddoomi^12,22^, Silvana I Albert^10^, Jonas Albinus^10^, Ilaria Alborelli^29^, Sonali Andani^9,22,31,36^, Per-Olof Attinger^14^, Marina Bacac^21^, Daniel Baumhoer^29^, Beatrice Beck-Schimmer^44^, Niko Beerenwinkel^7,22^, Christian Beisel^7^, Lara Bernasconi^32^, Anne Bertolini^12,22^, Bernd Bodenmiller^11,40^, Ximena Bonilla^9^, Lars Bosshard^12,22^, Byron Calgua^29^, Ruben Casanova^40^, Stéphane Chevrier^40^, Natalia Chicherova^12,22^, Ricardo Coelho^23^, Maya D’Costa^13^, Esther Danenberg^42^, Natalie R Davidson^9^, Monica-Andreea Dragan^7^, Reinhard Dummer^33^, Stefanie Engler^40^, Martin Erkens^19^, Katja Eschbach^7^, Cinzia Esposito^42^, André Fedier^23^, Pedro F Ferreira^7^, Joanna Ficek-Pascual^1,9,16,22,31^, Anja L Frei^36^, Bruno Frey^18^, Sandra Goetze^10^, Linda Grob^12,22^, Gabriele Gut^42^, Detlef Günther^8^, Pirmin Haeuptle^3^, Viola Heinzelmann-Schwarz^23,28^, Sylvia Herter^21^, Rene Holtackers^42^, Tamara Huesser^21^, Alexander Immer^9,17^, Anja Irmisch^33^, Francis Jacob^23^, Andrea Jacobs^40^, Tim M Jaeger^14^, Katharina Jahn^7^, Alva R James^9,22,31^, Philip M Jermann^29^, André Kahles^9,22,31^, Abdullah Kahraman^22,36^, Viktor H Koelzer^36,41^, Werner Kuebler^30^, Jack Kuipers^7,22^, Christian P Kunze^27^, Christian Kurzeder^26^, Kjong-Van Lehmann^2,4,9,15^, Mitchell Levesque^33^, Ulrike Lischetti^23^, Flavio C Lombardo^23^, Sebastian Lugert^13^, Gerd Maass^18^, Markus G Manz^35^, Philipp Markolin^9^, Martin Mehnert^10^, Julien Mena^5^, Julian M Metzler^34^, Nicola Miglino^35,41^, Emanuela S Milani^10^, Holger Moch^36^, Simone Muenst^29^, Riccardo Murri^43^, Charlotte KY Ng^29,39^, Stefan Nicolet^29^, Marta Nowak^36^, Monica Nunez Lopez^23^, Patrick GA Pedrioli^6^, Lucas Pelkmans^42^, Salvatore Piscuoglio^23,29^, Michael Prummer^12,22^, Prélot, Laurie^9,22,31^, Natalie Rimmer^23^, Mathilde Ritter^23^, Christian Rommel^19^, María L Rosano-González^12,22^, Gunnar Rätsch^1,6,9,22,31^, Natascha Santacroce^7^, Jacobo Sarabia del Castillo^42^, Ramona Schlenker^20^, Petra C Schwalie^19^, Severin Schwan^14^, Tobias Schär^7^, Gabriela Senti^32^, Wenguang Shao^10^, Franziska Singer^12,22^, Sujana Sivapatham^40^, Berend Snijder^5,22^, Bettina Sobottka^36^, Vipin T Sreedharan^12,22^, Stefan Stark^9,22,31^, Daniel J Stekhoven^12,22^, Tanmay Tanna^7,9^, Alexandre PA Theocharides^35^, Tinu M Thomas^9,22,31^, Markus Tolnay^29^, Vinko Tosevski^21^, Nora C Toussaint^12,22^, Mustafa A Tuncel^7,22^, Marina Tusup^33^, Audrey Van Drogen^10^, Marcus Vetter^25^, Tatjana Vlajnic^29^, Sandra Weber^32^, Walter P Weber^24^, Rebekka Wegmann^5^, Michael Weller^38^, Fabian Wendt^10^, Norbert Wey^36^, Andreas Wicki^35,41^, Mattheus HE Wildschut^5,35^, Bernd Wollscheid^10^, Shuqing Yu^12,22^, Johanna Ziegler^33^, Marc Zimmermann^9^, Martin Zoche^36^, Gregor Zuend^37^

^1^AI Center at ETH Zurich, Andreasstrasse 5, 8092 Zurich, Switzerland, ^2^Cancer Research Center Cologne-Essen, University Hospital Cologne, Cologne, Germany, ^3^Cantonal Hospital Baselland, Medical University Clinic, Rheinstrasse 26, 4410 Liestal, Switzerland, ^4^Center for Integrated Oncology Aachen (CIO-A), Aachen, Germany, ^5^ETH Zurich, Department of Biology, Institute of Molecular Systems Biology, Otto-Stern-Weg 3, 8093 Zurich, Switzerland, ^6^ETH Zurich, Department of Biology, Wolfgang-Pauli-Strasse 27, 8093 Zurich, Switzerland, ^7^ETH Zurich, Department of Biosystems Science and Engineering, Mattenstrasse 26, 4058 Basel, Switzerland, ^8^ETH Zurich, Department of Chemistry and Applied Biosciences, Vladimir-Prelog-Weg 1-5/10, 8093 Zurich, Switzerland, ^9^ETH Zurich, Department of Computer Science, Institute of Machine Learning, Universitätstrasse 6, 8092 Zurich, Switzerland, ^10^ETH Zurich, Department of Health Sciences and Technology, Otto-Stern-Weg 3, 8093 Zurich, Switzerland, ^11^ETH Zurich, Institute of Molecular Health Sciences, Otto-Stern-Weg 7, 8093 Zurich, Switzerland, ^12^ETH Zurich, NEXUS Personalized Health Technologies, Wagistrasse 18, 8952 Zurich, Switzerland, ^13^F. Hoffmann-La Roche Ltd, Grenzacherstrasse 124, 4070 Basel, Switzerland, ^14^F. Hoffmann-La Roche Ltd, Grenzacherstrasse 124, 4070 Basel, Switzerland,, ^15^Joint Research Center Computational Biomedicine, University Hospital RWTH Aachen, Aachen, Germany, ^16^Life Science Zurich Graduate School, Biomedicine PhD Program, Winterthurerstrasse 190, 8057 Zurich, Switzerland, ^17^Max Planck ETH Center for Learning Systems,, ^18^Roche Diagnostics GmbH, Nonnenwald 2, 82377 Penzberg, Germany, ^19^Roche Pharmaceutical Research and Early Development, Roche Innovation Center Basel, Grenzacherstrasse 124, 4070 Basel, Switzerland, ^20^Roche Pharmaceutical Research and Early Development, Roche Innovation Center Munich, Roche Diagnostics GmbH, Nonnenwald 2, 82377 Penzberg, Germany, ^21^Roche Pharmaceutical Research and Early Development, Roche Innovation Center Zurich, Wagistrasse 10, 8952 Schlieren, Switzerland, ^22^SIB Swiss Institute of Bioinformatics, Lausanne, Switzerland, ^23^University Hospital Basel and University of Basel, Department of Biomedicine, Hebelstrasse 20, 4031 Basel, Switzerland, ^24^University Hospital Basel and University of Basel, Department of Surgery, Brustzentrum, Spitalstrasse 21, 4031 Basel, Switzerland, ^25^University Hospital Basel, Brustzentrum & Tumorzentrum, Petersgraben 4, 4031 Basel, Switzerland, ^26^University Hospital Basel, Brustzentrum, Spitalstrasse 21, 4031 Basel, Switzerland, ^27^University Hospital Basel, Department of Information- and Communication Technology, Spitalstrasse 26, 4031 Basel, Switzerland, ^28^University Hospital Basel, Gynecological Cancer Center, Spitalstrasse 21, 4031 Basel, Switzerland, ^29^University Hospital Basel, Institute of Medical Genetics and Pathology, Schönbeinstrasse 40, 4031 Basel, Switzerland, ^30^University Hospital Basel, Spitalstrasse 21/Petersgraben 4, 4031 Basel, Switzerland, ^31^University Hospital Zurich, Biomedical Informatics, Schmelzbergstrasse 26, 8006 Zurich, Switzerland, ^32^University Hospital Zurich, Clinical Trials Center, Rämistrasse 100, 8091 Zurich, Switzerland, ^33^University Hospital Zurich, Department of Dermatology, Gloriastrasse 31, 8091 Zurich, Switzerland, ^34^University Hospital Zurich, Department of Gynecology, Frauenklinikstrasse 10, 8091 Zurich, Switzerland, ^35^University Hospital Zurich, Department of Medical Oncology and Hematology, Rämistrasse 100, 8091 Zurich, Switzerland, ^36^University Hospital Zurich, Department of Pathology and Molecular Pathology, Schmelzbergstrasse 12, 8091 Zurich, Switzerland, ^37^University Hospital Zurich, Rämistrasse 100, 8091 Zurich, Switzerland, ^38^University Hospital and University of Zurich, Department of Neurology, Frauenklinikstrasse 26, 8091 Zurich, Switzerland, ^39^University of Bern, Department of BioMedical Research, Murtenstrasse 35, 3008 Bern, Switzerland, ^40^University of Zurich, Department of Quantitative Biomedicine, Winterthurerstrasse 190, 8057 Zurich, Switzerland, ^41^University of Zurich, Faculty of Medicine, Zurich, Switzerland, ^42^University of Zurich, Institute of Molecular Life Sciences, Winterthurerstrasse 190, 8057 Zurich, Switzerland, ^43^University of Zurich, Services and Support for Science IT, Winterthurerstrasse 190, 8057 Zurich, Switzerland, ^44^University of Zurich, VP Medicine, Künstlergasse 15, 8001 Zurich, Switzerland

## Footnotes

1 https://portal.gdc.cancer.gov/

2 https://github.com/AMLab-Amsterdam/AttentionDeepMIL

3 https://github.com/mahmoodlab/SurvPath

4 https://github.com/mahmoodlab/MCAT

