## Supplementary Information for "Histopathology-based Protein Multiplex Generation using Deep Learning"

### S1 Supplement Information

#### S1.1 Benchmarking on Pixel-Aligned Datasets

To evaluate the performance of HistoPlexer on pixel-aligned datasets, we benchmark our method on two datasets: DeepLIIF singleplex immunohistochemistry (IHC) Ki67 and multiplex immunofluorescence (mIF) dataset [20] and DeepLIIF denovo mIF/mIHC stained dataset [14]. For simplicity, we denote these two datasets as DeepLIIF-A and DeepLIIF-B, respectively. Section S1.1.1 briefly describes the datasets and preprocessing, while Section S1.1.2 details the experimental results.

##### S1.1.1 DeepLIIF Datasets

**DeepLIIF-A.** The dataset comprises lung and bladder cancer tissues scanned with a ZEISS Axioscan scanner. For each case, images were acquired in five modalities: IHC, hematoxylin, and mIF for DAPI, Lap2, and Ki67, all from the same tissue sections. The training set includes tissues from three lung cancer patients (two males and one female, ages 45–57) and three bladder cancer patients (all males, ages 52–66), whereas the testing set contains tissues from two lung cancer patients (males, ages 48 and 55) and two bladder cancer patients (males, ages 61 and 68), with all patients being Caucasian. The dataset was downloaded from Zenodo[5].
**DeepLIIF-B.** The dataset comprises eight anonymized head-and-neck squamous cell carcinoma cases. For each case, nine regions-of-interest (ROIs) were selected—three from the tumor core, three from the tumor margin, and three from the adjacent stroma. Each ROI, sized at 1356×1012 pixels (0.343 $mm^2$ at 0.5 $\mu$m/pixel), was subdivided into 512×512 patches, yielding 268 co-registered image patches. Imaging protocols include both multiplex immunofluorescence (mIF) and multiplex immunohistochemistry (mIHC). The mIF staining utilized five markers: DAPI for nuclear staining; CD3, CD8, and FoxP3 as immune markers; and pan-cytokeratin (PanCK) as a tumor marker. Hematoxylin-stained images acquired via mIHC provided a complementary reference for tissue morphology. The dataset was downloaded from The Cancer Imaging Archive[6].
**Preprocessing**. All images were resized to 256×256 pixels to ensure uniform spatial dimensions and pixel intensities were normalized to the $[0, 1]$ range. For the immunofluorescence images, a multi-channel representation was created by stacking the individual markers' grayscale images along the channel dimension. We followed the official train-test split for the DeepLIIF-A dataset. For the DeepLIIF-B dataset, a random case-based split was performed by reserving two out of eight cases for testing, resulting in 199 training images and 69 testing images.

##### S1.1.2 Experimental Results

We evaluated HistoPlexer using hematoxylin images as input to predict the corresponding marker channels: DAPI, Lap2, and Ki67 for DeepLIIF-A, and DAPI, CD3, CD8, FoxP3, and PanCK for DeepLIIF-B. Experimental results, reported in Extended

[5] https://zenodo.org/records/4751737#.YKRTS0NKhH4
[6] https://www.cancerimagingarchive.net/collection/hnscc-mif-mihc-comparison/

Data Tables 1 and 2, show that HistoPlexer, operating in the multiplex (MP) setting, consistently outperforms benchmark methods in both perceptual similarity and pixel-level fidelity. Specifically, on DeepLIIF-A, HistoPlexer achieved an MS-SSIM of 0.656, a PSNR of 19.804, and an RMSE-SW of 0.072, while on DeepLIIF-B it attained an MS-SSIM of 0.704, a PSNR of 20.642, and an RMSE-SW of 0.092. These findings indicate that HistoPlexer not only improves performance on datasets with slice-to-slice variations but also excels on datasets with precise pixel-level alignment.

#### S1.2 Extended benchmarking on TuPro dataset

##### S1.2.1 Incorporating features from Computational Pathology Foundation Model

Recent advancements in computational pathology foundation models have greatly impacted the field, with many such models now publicly accessible. These models are predominantly utilized to derive rich representations of H&E stained images, which are then used in predictive downstream tasks. However, their potential for generative tasks involving HE images has not been extensively explored. In this study, we investigate the possibility of enhancing the generation of protein multiplexes by augmenting features derived from foundation models trained on H&E images.

For our experiments, we employed the UNI model [43], a widely adopted computational pathology foundation model [67]. UNI is based on the Vision Transformer (ViT-Large or ViT-L) architecture and is pretrained on approximately 100,000 HE whole slide images (WSIs), providing feature embeddings with a dimensionality of 1024. We integrated the UNI feature embeddings into the bottleneck layer of the U-Net translator model within the HistoPlexer framework, aiming to enrich the feature space and potentially enhance generative performance. To optimize computational efficiency, we pre-extracted the UNI feature embeddings and loaded them during training, resulting in a new model we refer to as HistoPlexer-FM.

As presented in Extended Data Table 3, HistoPlexer-FM performs comparably to the original HistoPlexer model on metrics such as MS-SSIM, PSNR and RMSE-SW. However, it exhibits significantly poorer performance on Mean Squared Error (MSE) for protein co-localization patterns, indicating a failure to capture biologically meaningful relationships between protein markers. This shortcoming may stem from the fact that UNI and similar foundation models are not trained for tasks involving protein multiplex generation requiring detailed spatial information of H&E images. This observation aligns with findings from [44], which demonstrate that end-to-end task-specific models often outperform those based on feature embeddings from UNI.

Through this exploration, we highlight the potential and limitations of integrating foundation model features in generative tasks, suggesting that while they can provide some benefits, task-specific training remains crucial for capturing complex biological interactions.

##### S1.2.2 Benchmarking with CycleGAN

We adopt the original CycleGAN implementation[7], utilizing its default configuration settings. In particular, we employ a generator based on a 9-block ResNet architecture. The L1 identity loss is disabled because it cannot be enforced when the input and output images possess different channel dimensions. Additionally, all images are resized to $256 \times 256$ pixels to comply with the required input dimensions.

As presented in Extended Data Table 3, we observe that CycleGAN consistently underperforms for all metrics. This observation is consistent with [12] that shows a better performance by Pix2pix over CycleGAN. It could be explained by the fact that the cycle consistency used in cycleGAN assumes a bijective mapping between the source and target domains, which does not hold for many stain translation tasks.

#### S1.3 Extended Interpretation of Results

##### S1.3.1 Biological Implications of Protein Co-localization Patterns

The co-localization patterns observed in Fig. 3 provide valuable insights into the simultaneous expression of specific protein markers within a tissue region. These patterns are particularly significant for pairs of markers of the same type, as they help elucidate the underlying biological processes. For example, a high Spearman correlation between CD3 and CD8a is expected, given that both are T cell markers, indicating active T cell infiltration. Similarly, the co-localization of MelanA and gp100, both tumor-associated antigens, underscores their role in identifying melanoma cells. The observed correlation between CD16 and HLA-DR suggests an activated yet impaired immune response, which could indicate the presence of regulatory or exhausted immune cells.

##### S1.3.2 Clinical Implications of Immune Phenotyping

Immune phenotyping, utilizing intratumoral and stromal CD8 density, aids in stratifying tumors into immune ”hot” and ”cold” categories, thus providing a framework for assessing immune activity within tumors. The immune phenotyping results, specifically the classification of tumors as immune ”hot” or ”cold,” have significant implications for therapeutic strategies, particularly immunotherapy. ”Hot” tumors, characterized by high CD8 T cell infiltration, are typically more responsive to immune checkpoint inhibitors, as they indicate a pre-existing anti-tumor immune response. These tumors may benefit from therapies that enhance T cell activity, such as PD-1/PD-L1 inhibitors. On the other hand, ”cold” tumors, with low CD8 infiltration, may require combination strategies to become more immunogenic. By tailoring immunotherapy strategies based on the immune landscape of tumors, clinicians can potentially improve patient outcomes and develop more personalized treatment regimens. Understanding these patterns not only aids in predicting responses to existing therapies but also guides the development of novel therapeutic approaches targeting specific immune-related pathways.

#### S1.4 Extended Figures and Tables

[7] https://github.com/junyanz/pytorch-CycleGAN-and-pix2pix

| | Method | MS-SSIM ↑ | PSNR ↑ | RMSE-SW ↓ |
|---|---|---|---|---|
| MP | Pix2pix [23] | 0.654 | 19.694 | 0.075 |
| | PyramidP2P [12] | 0.647 | 19.672 | 0.075 |
| | HistoPlexer | **0.656** | **19.804** | **0.072** |

**Extended Data Table 1**: Comparison of model performance on the DeepLIIF-A dataset in the multiplex (MP) setting using MS-SSIM, PSNR, and RMSE-SW. Arrows indicate whether higher (↑) or lower (↓) values are preferable.

| | Method | MS-SSIM ↑ | PSNR ↑ | RMSE-SW ↓ |
|---|---|---|---|---|
| MP | Pix2pix [23] | 0.675 | 20.132 | 0.097 |
| | PyramidP2P [12] | 0.638 | 18.726 | 0.114 |
| | HistoPlexer | **0.704** | **20.642** | **0.092** |

**Extended Data Table 2**: Comparison of model performance on the DeepLIIF-B dataset in the multiplex (MP) setting using MS-SSIM, PSNR, and RMSE-SW. Arrows indicate whether higher (↑) or lower (↓) values are preferable.

| | Method | MS-SSIM ↑ | PSNR ↑ | RMSE-SW ↓ | MSE-colocalization ↓ |
|---|---|---|---|---|---|
| MP | CycleGAN [27] | 0.130±0.031 | 4.585±0.713 | 0.587 ±0.047 | 0.1885 ±0.0273 |
| | Pix2pix [23] | 0.276±0.004 | 13.680±0.043 | 0.202 ±0.002 | <u>0.0070 ±0.0004</u> |
| | PyramidP2P [12] | 0.284±0.004 | 13.894±0.172 | 0.197±0.003 | 0.0081 ±0.0016 |
| | HistoPlexer | **0.300±0.003** | <u>14.162±0.076</u> | <u>0.195±0.001</u> | **0.0068 ±0.0005** |
| | HistoPlexer-FM | <u>0.292±0.005</u> | **14.552±0.104** | **0.189±0.001** | 0.0129 ±0.0015 |
| SP | CycleGAN [27] | 0.138±0.007 | 5.175±0.090 | 0.548 ±0.012 | 0.3084 ±0.0519 |
| | Pix2pix [23] | 0.260±0.004 | 13.015±0.009 | 0.218±0.001 | 0.0334 ±0.0011 |
| | PyramidP2P [12] | 0.264±0.015 | 13.216±0.483 | 0.214±0.011 | **0.0299 ±0.0056** |
| | HistoPlexer | **0.279±0.002** | **13.354±0.038** | **0.210±0.001** | <u>0.0302 ±0.0012</u> |
| | HistoPlexer-FM | <u>0.267±0.014</u> | <u>13.248±0.206</u> | <u>0.213±0.002</u> | 0.0514 ±0.0233 |

**Extended Data Table 3**: Comparison of Model Performance against benchmarks using MS-SSIM, PSNR, RMSE-SW and MSE for co-localization patterns for multiplex (MP) and singleplex (SP) settings. ↑ arrow indicates higher values are better. ↓ arrow indicates higher values are better.

| Protein | MSSSIM ↑ | PSNR ↑ | RMSE-SW ↓ |
|---|---|---|---|
| CD16 | 0.221±0.006 | 12.419±0.086 | 0.185±0.003 |
| CD20 | 0.510±0.017 | 19.340±0.321 | 0.104±0.003 |
| CD3 | 0.321±0.005 | 15.284±0.118 | 0.147±0.002 |
| CD31 | 0.685±0.007 | 24.546±0.394 | 0.054±0.001 |
| CD8a | 0.405±0.007 | 16.700±0.142 | 0.116±0.001 |
| HLA-ABC | 0.077±0.005 | 9.273±0.081 | 0.327±0.003 |
| HLA-DR | 0.224±0.010 | 12.530±0.136 | 0.188±0.004 |
| MelanA | 0.246±0.005 | 11.673±0.157 | 0.253±0.003 |
| S100 | 0.217±0.007 | 10.649±0.056 | 0.281±0.003 |
| SOX10 | 0.211±0.010 | 12.606±0.341 | 0.217±0.009 |
| gp100 | 0.179±0.005 | 10.761±0.143 | 0.274±0.002 |

**Extended Data Table 4**: Performance of HistoPlexer using MSSSIM, PSNR, RMSE-SM metrics for each marker, presented as mean±standard deviation.

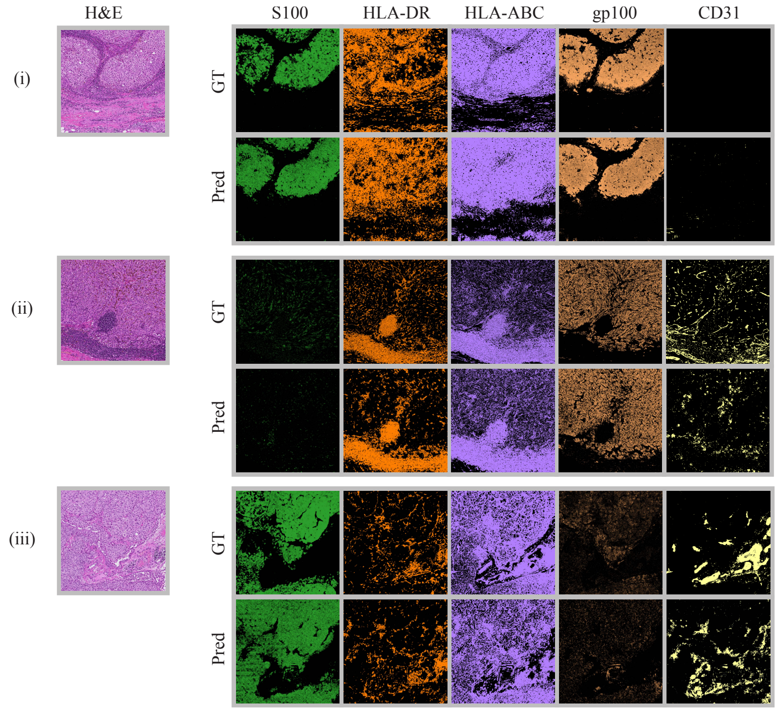


**Extended Data Fig. 1**: Qualitative RoI-level assesment of HistoPlexer. H&E (first column) and expression profiles of individual markers: S100, HLA-DR, HLA-ABC, gp100 and CD31 (from second to last column). Top row: ground-truth (GT) expression profiles; bottom row: predicted (pred) expression profiles.

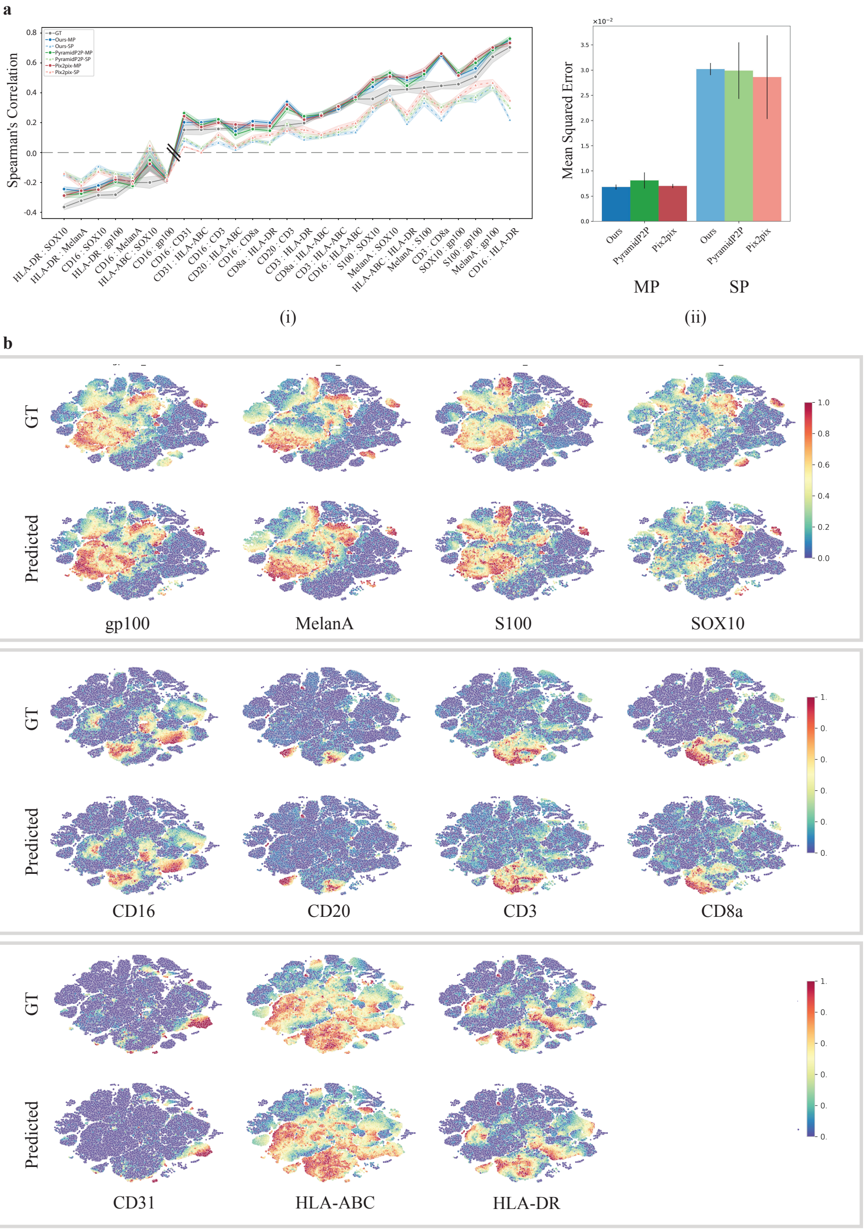


**Extended Data Fig. 2**: **a**(i) Spearman's correlation coefficients between protein pairs, comparing the ground truth (GT) with both singleplexd (SP) and multiplexed (MP) predictions of the HistoPlexer. **a**(ii) Mean squared error between the GT and predicted Spearman's correlation coefficients, comparing the SP and MP predictions of the HistoPlexer. **b** Joint t-SNE visualization of protein co-localization patterns.

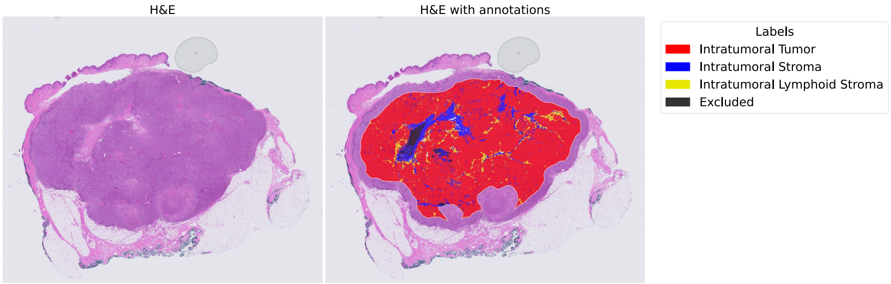


**Extended Data Fig. 3**: Region annotation for Immune phenotyping. H&E WSI (left) and H&E WSI with outline of Tumor Center compartment and annotated regions (right).

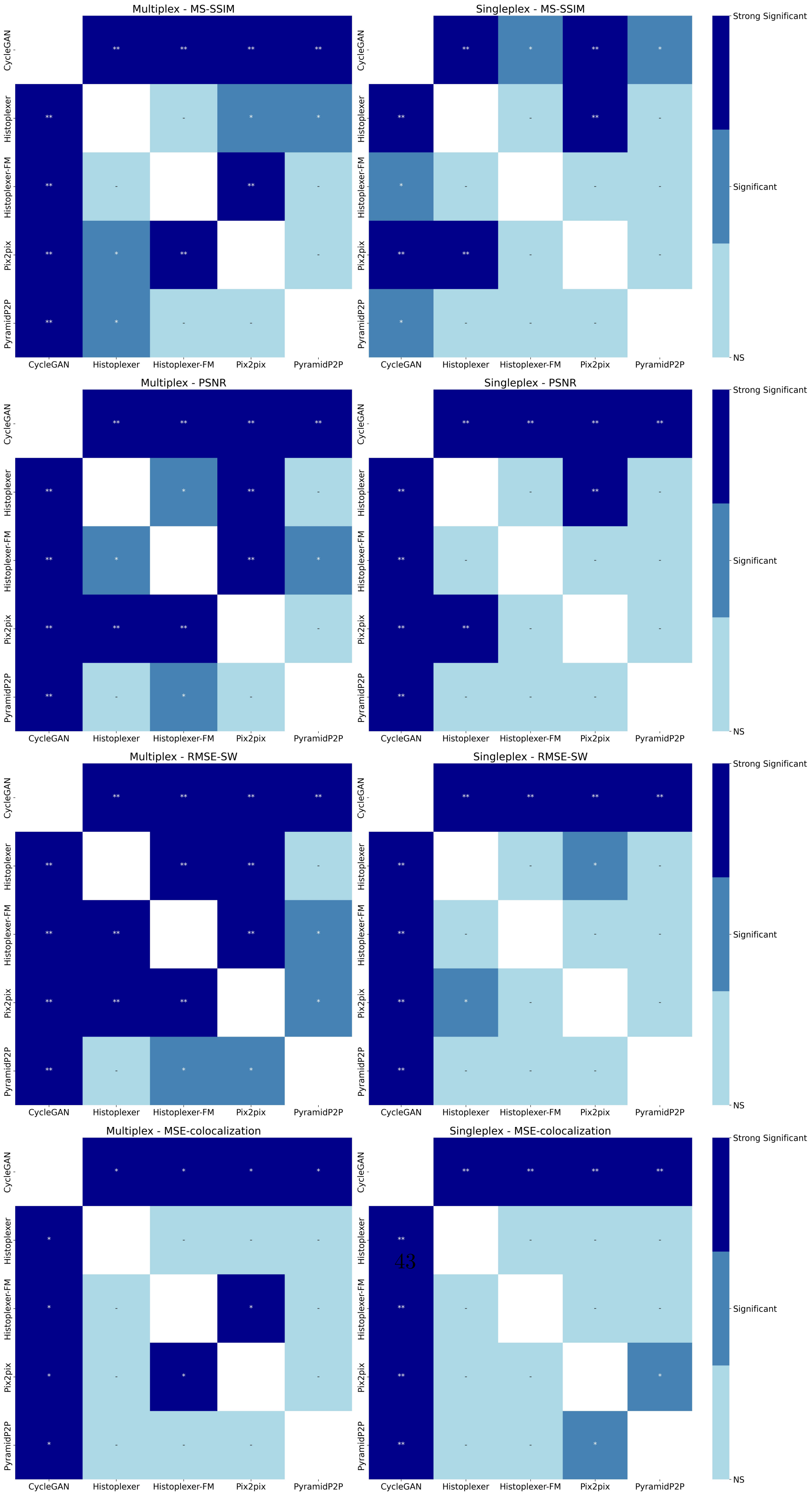


**Extended Data Fig. 4**: Paired t-test: Comparison of significance levels between methods in multiplex (left) and singleplex (right) settings for MS-SSIM (row 1), PSNR (row 2), RMSE-SW (row 3) and MSE-colocalization (row 4) metrics. Colorbar indicating significance levels: Not Significant (NS) (p≥0.05), Significant (p<0.05), and Strong Significant (p<0.01).
